# Risk factor targeting for vaccine prioritization during the COVID-19 pandemic

**DOI:** 10.1101/2021.03.04.21251264

**Authors:** Lloyd A. C. Chapman, Poojan Shukla, Isabel Rodríguez-Barraquer, Priya B. Shete, Tomás M. León, Kirsten Bibbins-Domingo, George W. Rutherford, Robert Schechter, Nathan C. Lo

## Abstract

A key public health question during any disease outbreak when limited vaccine is available is who should be prioritized for early vaccination. Most vaccine prioritization analyses only consider variation in risk of infection and death by a single risk factor, such as age. We provide a more granular approach with stratification by demographics, risk factors, and location. We use this approach to compare the impact of different COVID-19 vaccine prioritization strategies on COVID-19 cases, deaths and disability-adjusted life years (DALYs) over the first 6 months of vaccine rollout, using California as a case example. We estimate the proportion of cases, deaths and DALYs averted relative to no vaccination for strategies prioritizing vaccination by a single risk factor and by multiple risk factors (e.g. age, location). We find that age-based targeting averts the most deaths (62% for 5 million individuals vaccinated) and DALYs (38%) of strategies targeting by a single risk factor and targeting essential workers averts the least deaths (31%) and DALYs (24%) over the first 6 months of rollout. However, targeting by two or more risk factors simultaneously averts up to 40% more DALYs. Our findings highlight the potential value of multiple-risk-factor targeting of vaccination against COVID-19 and other infectious diseases.

## Introduction

The COVID-19 pandemic has caused significant morbidity and mortality, alongside social and economic disruption globally ^1–3^. SARS-CoV-2, the acute respiratory virus causing the clinical disease COVID-19, was first reported to the World Health Organization on December 31, 2019, as an outbreak of a novel pneumonia in Wuhan, China. As of December 30, 2021, COVID-19 has caused over 50 million reported cases and 800,000 deaths in the United States alone and significant burden in low and middle-income countries ^4^. The principal public health strategies have focused on stay-at-home orders and social distancing measures to minimize cases and transmission, and vaccination, once vaccines became widely available ^1–3^.

In December 2020, the United States Food and Drug Administration issued Emergency Use Authorization for the Pfizer/BioNTech BNT162b2-mRNA and Moderna mRNA-1273 vaccines that prevent or reduce severity of COVID-19 ^5–8^. Analyses of clinical trials and evidence from early vaccine roll-out demonstrated that these vaccines are both around 90-95% efficacious at preventing clinical disease ^8,9^.

A key public health question given availability of a COVID-19 vaccine has been how to optimally and equitably allocate the limited initial doses of the vaccine across risk groups. Key considerations have included whether to focus on reducing deaths, overall cases and transmission, or composite measures, such as disability-adjusted life years (DALYs) alongside equity considerations. Many unique epidemiologic risk groups are at higher risk of infection and/or poor outcome from COVID-19 and could be prioritized such as the elderly, frontline workers, and members of vulnerable populations such as incarcerated adults and persons experiencing homelessness. To aid decision makers, prioritization frameworks were proposed by the US Centers for Disease Control and Prevention Advisory Committee on Immunization Practices (CDC ACIP), the National Academies of Sciences, Engineering, and Medicine (NASEM), as well as the World Health Organization’s Strategic Advisory Group of Experts on Immunization (WHO SAGE) ^10–15^. These frameworks broadly agreed that the scope of maximizing human well-being involves not only reducing deaths, but also supporting workers who are vital to critical industries. Thus, while minimizing mortality should be an important goal for vaccine prioritization strategies, mitigating transmission and morbidity, particularly among essential workers, may also be a key consideration. Furthermore, the proposed frameworks highlighted the importance of not exacerbating health inequities through vaccine prioritization, as African American, Latinx and other minority communities are overrepresented among cases, hospitalizations, deaths, and the essential worker population in the US, while being underrepresented among individuals above the age of 65 ^10^.

Vaccine distribution frameworks also identified key populations and strategies for prioritization. However, data on the impact of prioritization of different risk groups was relatively limited. Existing analyses of different vaccine prioritization strategies have focused on targeting of vaccinations by age and potentially occupation ^16–22^, and generally agree that targeting older individuals first is the best strategy to minimize COVID-19 health burden when vaccine supply is limited ^23^. However, most have not considered the potential benefit of targeting vaccinations by multiple risk factors at the same time as we do here. While age is the main risk factor for severe disease and death from COVID-19, differences in risk of infection and death with other factors, such as location and sex, can potentially offset or exacerbate the increase in death risk with increasing age.

Although over 50% of the world’s population has now received at least one dose of a COVID-19 vaccine, coverage remains very low (<10%) in many low-income countries, so there is still some scope for optimizing targeting of early vaccinations in these countries. COVID-19 also provides a test case for assessing the potential benefit of targeting vaccination by multiple risk factors simultaneously for future epidemics. We developed a simulation model to evaluate targeting limited initial vaccine doses by multiple risk factors and applied this to prioritization of early COVID-19 vaccination. We used the case example of California in the United States that experienced a steep increase in cases and deaths in late 2020 prior to the arrival of vaccinations ^4^.

## Methods

### Model structure

We developed a simulation model of COVID-19 risk in the population of California to simulate the epidemiologic impact of different COVID-19 vaccine prioritization strategies. We selected California based on the availability of detailed person-level data on clinical cases, demographic variables, and risk factors. We first simulated the California population (N=39.1 million ^24^) by assigning the following demographic characteristics to each person: age, sex, race, ethnicity, county of residence (see Supplementary Information for variable definitions), using 5-year estimates from the 2018 American Community Survey ^24^. We then incorporated a ‘special population’ status for certain individuals, explicitly including healthcare workers, skilled nursing facility (SNF) residents, assisted living facility (ALF) residents, prisoners, education workers, persons experiencing homelessness, frontline essential workers, and non-frontline essential workers (see Supplementary Information for details). These were based on the NASEM prioritization framework as well as CDC ACIP definitions for frontline and non-frontline essential workers and represent a stratification of infection and mortality risk differentiated from the general population ^10,25^. We used data from the cross-sectional, geotagged California Health Interview Survey (CHIS) ^26^ to estimate comorbidity status in the Californian population, including binary status for asthma, diabetes, heart disease, heart failure, hypertension, obesity, and tobacco use.

### Calibration and validation

We calibrated the model to estimate risk of COVID-19 mortality using data provided by the California Department of Public Health on 28,175 COVID-19 deaths from February 5, 2020 to December 30, 2020. The de-identified data included information on age, sex, race, ethnicity, county of residence, date of death, and date of laboratory diagnosis (see Supplementary Information for variable definitions) for each patient. We applied a Poisson regression to estimate the relationship between risk of death from COVID-19 and age, sex, race/ethnicity, and county of residence. COVID-19 death risk estimates were further adjusted using literature estimates of relative risk of infection or death for each special population and of death given infection for each comorbidity (see Supplementary Information). We used data on numbers of COVID-19 deaths in California by age and long-term care facility residency status to estimate the cumulative number of non-long-term-care-facility deaths in each demographic risk group. We then estimated cumulative infections in each demographic risk group of individuals not living in long-term care facilities by dividing cumulative deaths by a published ensemble estimate of the age- and sex-specific infection fatality rate (IFR) ^27^ for non-long-term-care facility residents. Infections among long-term care facility residents were estimated by dividing by the age- and sex-specific IFR multiplied by a factor representing the frailty of long-term care facility residents relative to the general population, which we assumed is 3 ^27^. Cumulative clinical cases were then estimated by multiplying cumulative infections by an estimate of the age-dependent clinical fraction obtained from the literature ^28^.

The IFR was recalibrated to California by multiplying it by a factor such that the estimated total clinical cases from the model matched the total confirmed cases in the California Department of Public Health data, and estimated total infections agreed with seroprevalence estimates for California from the CDC’s nationwide commercial laboratory serosurveys ^29^ (see Supplementary Information). For the forward simulations, the number of previously infected individuals in each demographic-special-population-comorbidity risk group was drawn from a binomial distribution whose parameters were the group population size and the probability of previous infection based on the estimated number of infections. The cumulative number of deaths in each risk group over the prediction time horizon was then simulated from a binomial distribution whose parameters were the number of initially uninfected individuals in the group and their cumulative probability of death over the prediction horizon. Cumulative numbers of infections and clinical cases in each risk group were then calculated from the predicted deaths using the recalibrated IFR and clinical fraction as described above. To check the epidemiologic simulation model, we re-fitted it to data on COVID-19 deaths up to September 30, 2020; predicted deaths for October 1 to December 30, 2020, via 1000 simulations of the refitted model; and compared the simulated deaths against observed deaths in the data for October-December 2020. We note, however, that the goal of this study was not forecasting of COVID-19 cases and deaths.

### Study outcomes

The study outcomes included infections, clinical cases, deaths, and DALYs from COVID-19. SARS-CoV-2 infections were defined as individuals positive for SARS-CoV-2 RNA with or without clinical symptoms. A clinical COVID-19 case was defined as an individual positive for SARS-CoV-2 RNA (either confirmed by a diagnostic test, such as nucleic acid amplification, or not) with clinical symptoms. We defined COVID-19 deaths as those deemed directly attributable to COVID-19. DALYs were estimated as a combination of years of life lost from premature death due to COVID-19 and disability associated with the acute infection period (see Supplementary Information). We estimated mean years of life lost per death in each risk group by averaging estimates of years of life remaining for individuals in the group given their age, sex and race/ethnicity from US life tables ^30^. We applied disability weights for the acute infection period based on weights for episodes of “mild”, “moderate” and “severe” acute illness from previous literature ^31^, and did not include long-term sequelae of illness.

### Simulation of vaccination strategies

We evaluated five strategies for vaccine allocation that targeted vaccination by a single risk factor: (i) random allocation (lottery), (ii) special population targeting, (iii) age-based targeting, (iv) essential worker targeting, and (v) comorbidity targeting. Under random allocation, vaccinations were randomly distributed among individuals aged ≥20 years (younger individuals were not included as the Pfizer and Moderna vaccines were initially only authorized for use in individuals ≥16 years of age and ≥18 years of age ^5,6^). Under all other prioritization strategies, subgroups among individuals ≥20 years were ranked by estimated average DALY risk per person (see Supplementary Information), and vaccines allocated in order of decreasing risk. Under special population targeting, vaccines were allocated to incarcerated adults, then education workers, and then persons experiencing homelessness (based on highest risk in these groups). Under age-based targeting, vaccines were allocated from the oldest age group (≥80 years) to the youngest (20-to-29-year-olds) in 10-year age bins due to the increasing risk of death with older age. Under essential worker targeting, vaccines were first allocated to frontline essential workers due to their higher infection risk, and then to non-frontline essential workers. Frontline and non-frontline essential workers, excluding healthcare and education workers who were included as separate groups, were defined as per occupational categories provided by the US CDC ^25^. Under comorbidity targeting, individuals with any comorbidities (see Supplementary Information for list) were prioritized over those with no comorbidities due to their higher death risk. We then evaluated strategies that targeted vaccination simultaneously by two risk factors: age-and-county targeting and age-and-special-population targeting, and a strategy in which vaccinations were targeted by all risk factors simultaneously, i.e. all demographic-special-population-comorbidity risk groups were allocated vaccinations in descending order of estimated DALY risk per person. For all strategies, we assumed all healthcare workers and residents of long-term care facilities, i.e. SNFs and ALFs, were vaccinated first based on the US CDC Phase 1a decision ^12^. The prioritization strategies considered were based on published frameworks for COVID-19 vaccine allocation and discussion with policymakers ^10,12,13,25^. COVID-19 deaths, clinical cases and infections over 6 months from the start of 2021 in each demographic-special-population-comorbidity risk group were simulated 1000 times for each strategy using the fully-adjusted risk model described above, but with different levels of protection against infection, clinical disease and death provided by vaccination or previous infection accounted for in the calculation of infections and clinical cases (see Supplementary Information). Uncertainty in the parameter estimates of the risk model and relative risks of infection for the special populations was accounted for by sampling their values from truncated normal distributions in each simulation (see Supplementary Information), and approximate 95% uncertainty intervals for the cumulative numbers of infections, cases, deaths, and DALYs were calculated as the 2.5%-97.5% quantiles of the predicted distributions. We modeled varying levels of vaccine availability, including 2 million, 5 million, and 10 million persons being vaccinated (corresponding to 5%, 13%, and 26% of the California population, respectively), taking 5 million persons vaccinated as the base case scenario. We assumed that protection against death, clinical disease and infection from vaccination were initially 95%, 90% and 80% ^32^, but that vaccination had no impact on transmission. We accounted for waning of immunity by assuming that vaccine-derived protection against infection and death declined exponentially over time at rates based on published estimates ^33^ (see Supplementary Information). We also assumed vaccination and onset of protection was instantaneous at the start of the simulation and vaccine adherence was 100%. Deaths from reinfections were modelled by assuming that previous infection conferred the same degree of protection as two-dose vaccination and that this protection waned at the same rate.

### Sensitivity analyses

We repeated the analysis for different vaccine efficacy profiles: 60% age-stable efficacy against death, given other SARS-CoV-2 vaccinations in use globally with lower efficacy; and 20% age-stable efficacy, to assess the potential impact of a very-low-efficacy vaccine. We also reran the analysis using quality-adjusted life years (QALYs) instead of DALYs -- prioritizing vaccinations by maximum risk of QALY loss -- since QALYs can provide different insights and different conclusions from DALYs in health economic analyses^34^.

### Ethics approval and consent to participate

This study was deemed eligible for expedited review as research involving data collected solely for non-research purposes and was approved by the University of California, San Francisco Institutional Review Board. All research was performed in accordance with relevant guidelines and regulations. The Institutional Review Board approved a waiver of informed consent for all participants on the basis that the research posed no more than a minimal risk to participants and would not adversely affect their rights or welfare.

## Results

### Demographic patterns of cases and deaths, model calibration, and validation

As of December 30, 2020, 2,215,972 confirmed COVID-19 cases and 28,175 COVID-19 deaths had occurred in California. The age distribution of confirmed cases was skewed towards younger individuals, with a peak in cumulative incidence in 20-to-29-year-olds, while the cumulative incidence of death increased exponentially with age, peaking at just under 0.8% in individuals aged ≥80 years (Figure 1). Cumulative COVID-19 case incidence and COVID-19-attributable death incidence varied considerably across counties from 2.8% to 12.6% and 0.02% to 0.2% respectively. Cumulative case incidence was slightly higher among females (6.0%) than males (5.8%), but this trend was reversed for deaths (cumulative incidence = 0.06% in females and 0.08% in males). Absolute numbers of both COVID-19 cases and deaths were highest among Hispanic/Latinx individuals, while cumulative incidence of death was highest among non-Hispanic Black individuals (0.09%). The demographic and geographic patterns in the death hazard ratio estimates from the model were similar to those in the cumulative incidence of death, with age by far the strongest risk factor, although death risk was highest for Hispanic/Latinx individuals (Figure S2).

**Figure 1.**
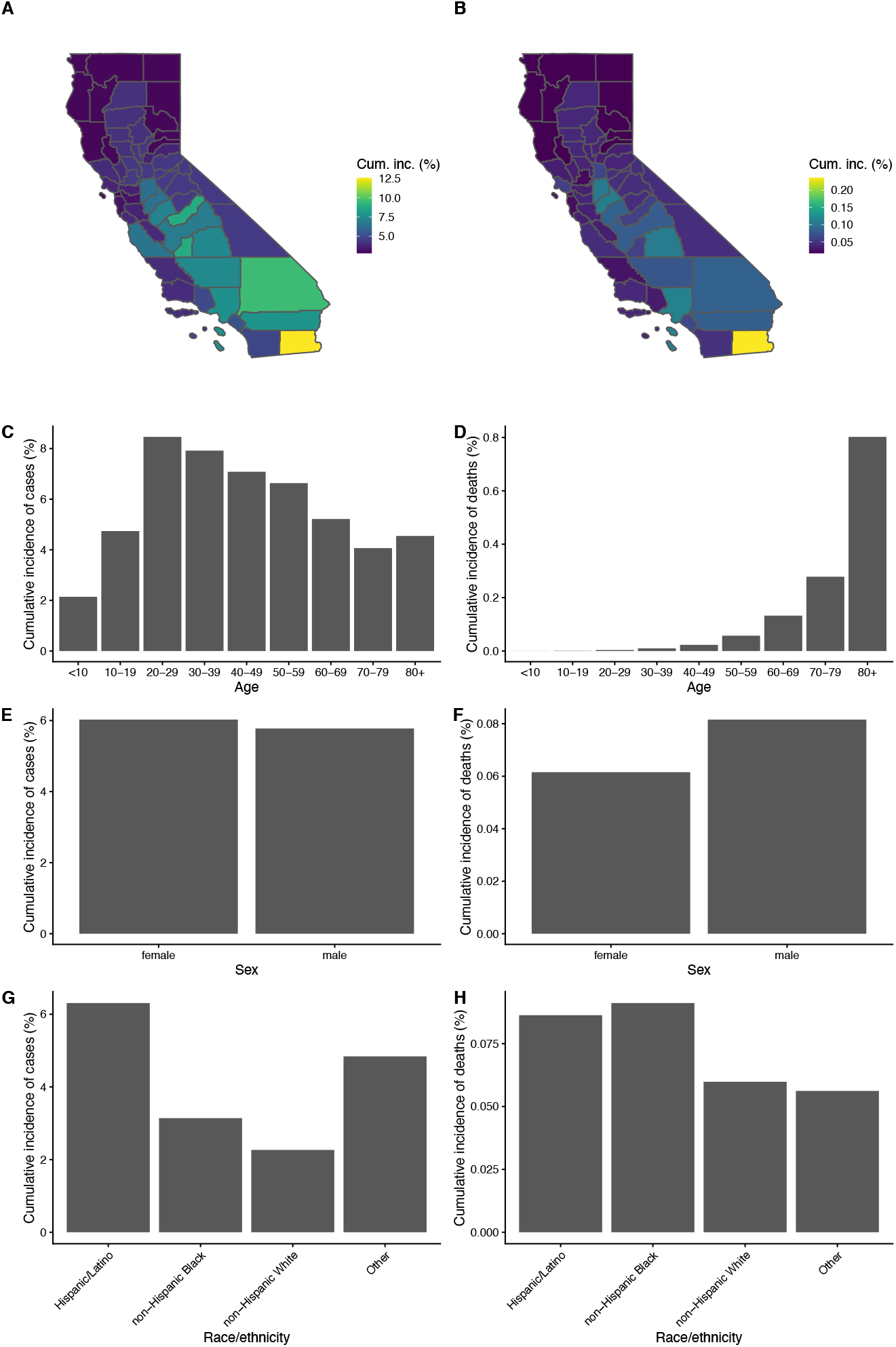
Cumulative incidence of confirmed COVID-19 cases (left column) and deaths (right column) in California up to December 30, 2020, by key demographic factors (rows). Cumulative incidence by (A-B) county of residence, (C-D) age, (E-F) sex, and (G-H) race/ethnicity. Data from California Department of Public Health. Counties with population <250,000 (except Imperial) were combined into a single region by their economic region, San Benito County was combined with Santa Cruz County, and Napa County was combined with Sonoma County. Plotted cumulative incidence for these counties represents the cumulative incidence of the combined region.

Calibration of the model to the state-level cumulative number of confirmed cases yielded age distributions of cases and infections close to those of confirmed cases in the California Department of Public Health data and infections estimated from CDC state-level seroprevalence data ^29^ (Figure 2). There was also close correspondence between the model estimates and observed data in terms of patterns of cases by county, sex, and race/ethnicity (Figure S1). Predictions from fitting the model to data up to the end of September 2020 accurately captured the geographic, age, sex, and race/ethnicity patterns of observed deaths for October to December 2020, although they underestimated the number of deaths to some extent (Figure S4).

**Figure 2.**
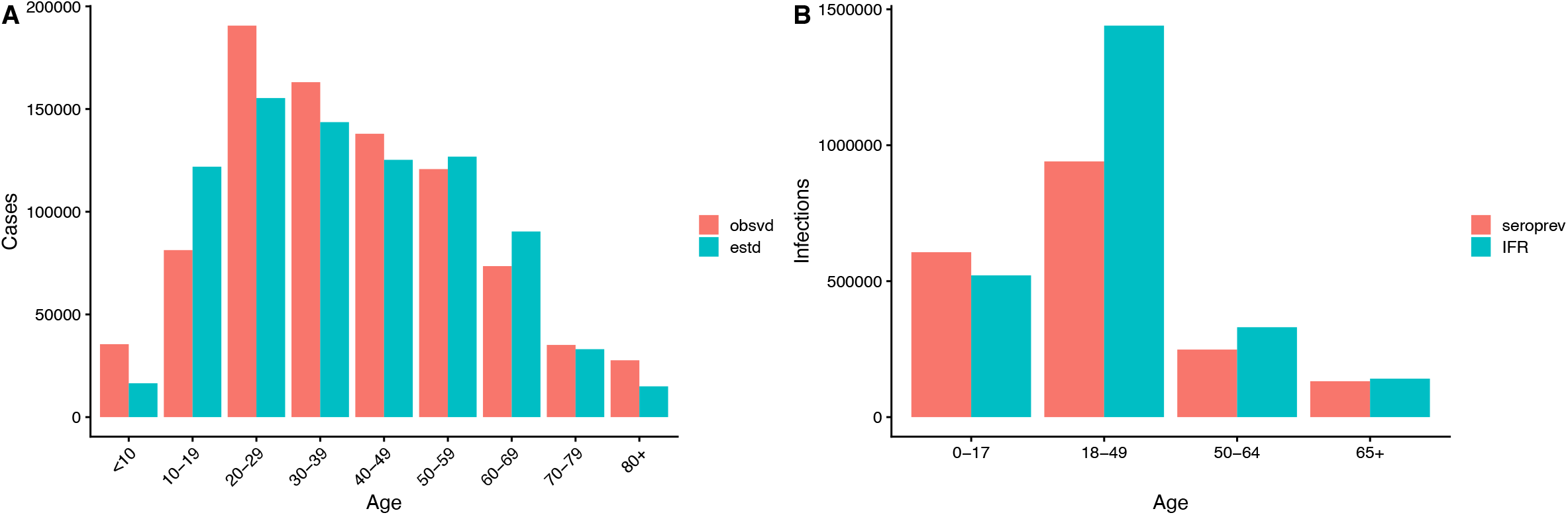
Model calibration. (A) Age distributions of cumulative numbers of confirmed cases in California Department of Public Health data (red) and clinical cases estimated from the model (turquoise) up to October 22, 2020. (B) Age distributions of infections estimated from seroprevalence estimates for California up to October 8-13, 2020, from CDC nationwide commercial lab seroprevalence data (red) ^29^ and estimated from the model using the infection fatality rate (IFR) from ^27^ recalibrated for California such that the total numbers of cases in the distributions in (A) match.

### Impact of vaccine prioritization strategies

Under the counterfactual scenario of no vaccination, the simulated total disease burden from COVID-19 in California in the first six months of 2021 according to the model was 211,000 DALYs (95% UI 206,000-215,000), resulting from 1.7 million infections (95% UI 1.5-2.0 million), 610,000 clinical cases (95% UI 560,000-670,000), and 14,100 deaths (95% UI 13,900-14,300). In terms of averting DALYs, age-targeting performed the best of the strategies targeting by a single risk factor, followed by comorbidity targeting, then special population targeting and essential worker targeting (Table 1 and Figure 3C). In the base case scenario (5 million vaccinations), age-targeting averted 81,000 DALYs (95% UI 78,000-83,000), amounting to 38% (95% UI 37-39%) of the simulated overall DALY burden without any vaccination. Comorbidity-targeting averted 59,000 DALYs (95% UI 56,000-62,000; 28% of total burden). Prioritizing special populations and essential workers averted 52,000 DALYs (95% UI 49,000-55,000; 25% of total) and 50,000 DALYs (95% UI 48,000-53,000; 24% of total) respectively, slightly fewer than random allocation (54,000 DALYS, 95% UI 51,000-56,000; 25% of total) due to a smaller proportion of older individuals being vaccinated under these strategies.

**Table 1.**
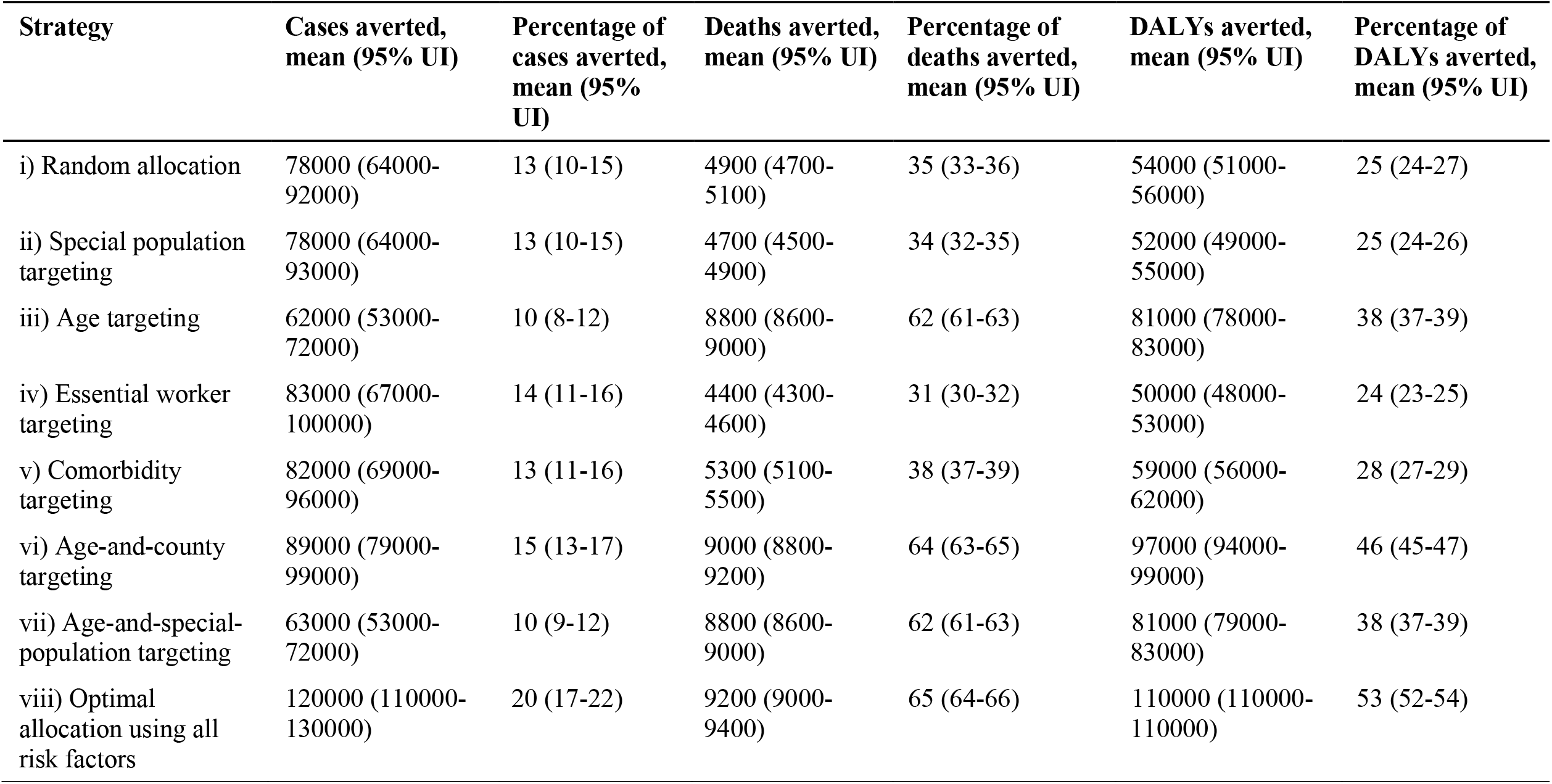
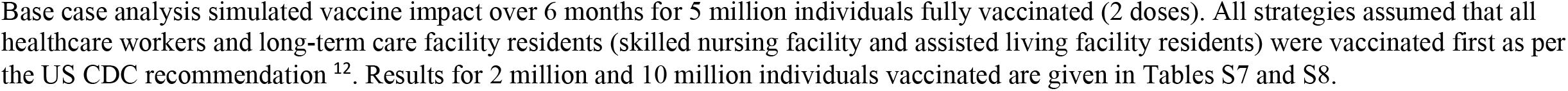
**Simulated impact of different COVID-19 vaccine prioritization strategies in California on numbers of averted COVID-19 clinical cases, deaths and DALYs for 5 million people vaccinated**

| Strategy | Cases averted,<br>mean (95% UI) | Percentage of<br>cases averted,<br>mean (95%<br>UI) | Deaths averted,<br>mean (95% UI) | Percentage of<br>deaths averted,<br>mean (95% UI) | DALYs averted,<br>mean (95% UI) | Percentage of<br>DALYs averted,<br>mean (95% UI) |
| --- | --- | --- | --- | --- | --- | --- |
| i) Random allocation | 78000 (64000-92000) | 13 (10-15) | 4900 (4700-5100) | 35 (33-36) | 54000 (51000-56000) | 25 (24-27) |
| ii) Special population targeting | 78000 (64000-93000) | 13 (10-15) | 4700 (4500-4900) | 34 (32-35) | 52000 (49000-55000) | 25 (24-26) |
| iii) Age targeting | 62000 (53000-72000) | 10 (8-12) | 8800 (8600-9000) | 62 (61-63) | 81000 (78000-83000) | 38 (37-39) |
| iv) Essential worker targeting | 83000 (67000-100000) | 14 (11-16) | 4400 (4300-4600) | 31 (30-32) | 50000 (48000-53000) | 24 (23-25) |
| v) Comorbidity targeting | 82000 (69000-96000) | 13 (11-16) | 5300 (5100-5500) | 38 (37-39) | 59000 (56000-62000) | 28 (27-29) |
| vi) Age-and-county targeting | 89000 (79000-99000) | 15 (13-17) | 9000 (8800-9200) | 64 (63-65) | 97000 (94000-99000) | 46 (45-47) |
| vii) Age-and-special-population targeting | 63000 (53000-72000) | 10 (9-12) | 8800 (8600-9000) | 62 (61-63) | 81000 (79000-83000) | 38 (37-39) |
| viii) Optimal allocation using all risk factors | 120000 (110000-130000) | 20 (17-22) | 9200 (9000-9400) | 65 (64-66) | 110000 (110000-110000) | 53 (52-54) |

Base case analysis simulated vaccine impact over 6 months for 5 million individuals fully vaccinated (2 doses). All strategies assumed that all healthcare workers and long-term care facility residents (skilled nursing facility and assisted living facility residents) were vaccinated first as per the US CDC recommendation <sup>12</sup>. Results for 2 million and 10 million individuals vaccinated are given in Tables S7 and S8.

**Figure 3.**
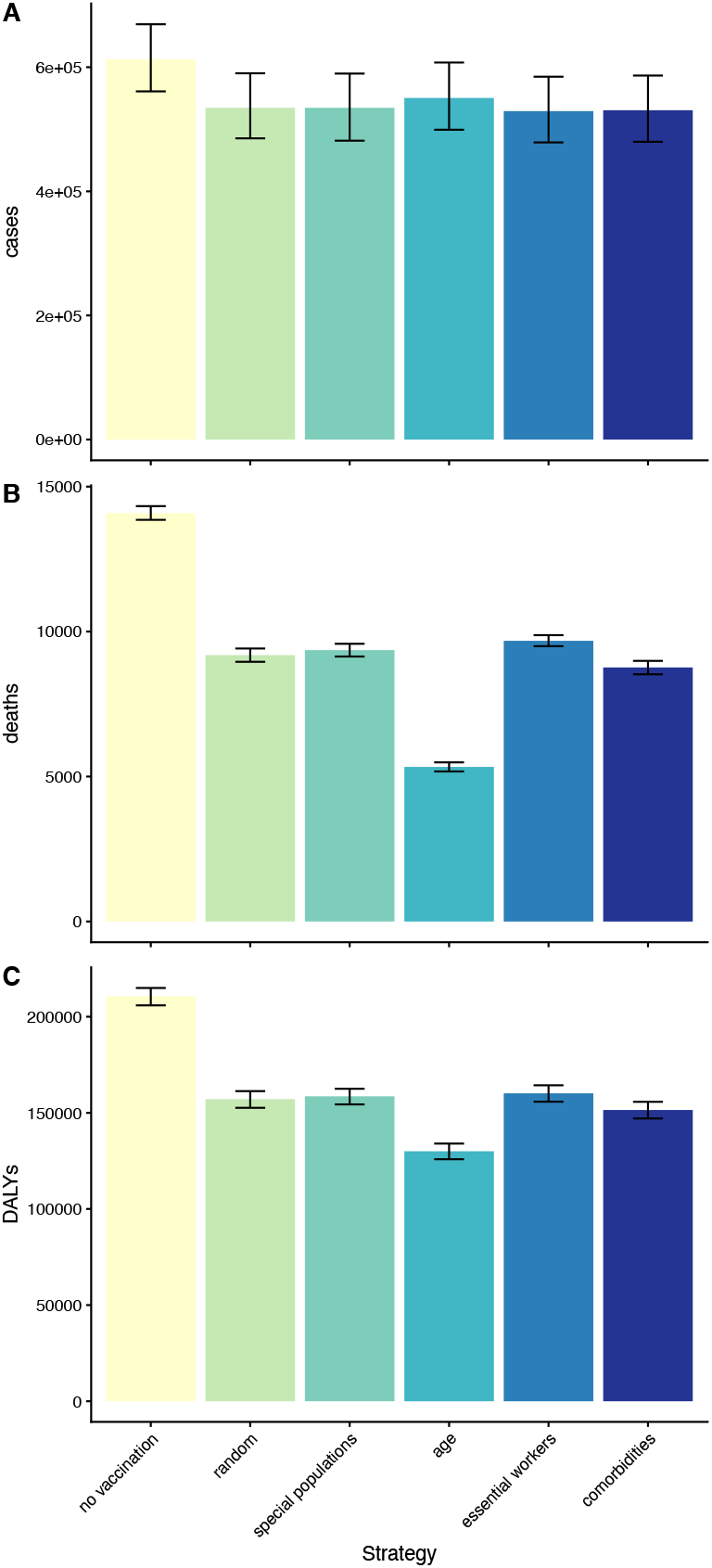
Simulated impact of different COVID-19 vaccine prioritization strategies in California on numbers of COVID-19 clinical cases, deaths and DALYs for 5 million people vaccinated. Simulated (A) clinical cases, (B) deaths and (C) DALYs in California in the first 6 months of 2021. Five different prioritization strategies were considered: (i) random allocation, (ii) targeting special populations (prisoners, education workers, people experiencing homelessness), (iii) age targeting, (iv) targeting essential workers, (v) targeting individuals with comorbidities. All strategies assumed that all healthcare workers and long-term care facility residents were vaccinated first as per the US CDC recommendation ^12^. Vaccine efficacies against clinical disease and death were assumed to be initially 90% and 95% and to wane gradually over time (see Supplementary Information). Bars show mean estimates across 1000 simulations, error bars show 95% uncertainty intervals from stochastic uncertainty and parameter uncertainty. Results for 2 million and 10 million individuals vaccinated are shown in Figure S3.

The order of performance of the strategies was the same for deaths averted: age-targeting averted most deaths, then comorbidity-targeting, then special population-targeting and essential worker-targeting (Table 1 and Figure 3B). Age targeting averted 8,800 deaths (95% UI 8,600-9,000; 62% of total deaths). Comorbidity-targeting, special population-targeting and essential worker-targeting averted 5,300 deaths (95% UI 5,100-5,500; 38% of total), 4,700 deaths (95% UI 4,500-4,900; 34% of total) and 4,400 deaths (95% UI 4,300-4,600; 31% of total), respectively.

In terms of clinical cases averted, essential-worker-targeting performed best, followed by comorbidity-targeting, special-population-targeting and age-targeting (Table 1 and Figure 3A). Essential-worker-targeting averted 83,000 cases (95% UI 67,000-100,000, 14% of total). Comorbidity-targeting and special-population-targeting averted 82,000 cases (95% UI 69,000-96,000; 13% of total cases) and 78,000 cases (95% UI 64,000-93,000; 13% of total) respectively. Age-targeting averted 62,000 cases (95% UI 53,000-72,000; 10% of total), fewer even than random allocation (78,000, 95% UI 64,000-92,000; 13% of total).

The main study findings were found to be robust across different vaccine availabilities (2-10 million people vaccinated) (Tables S7 and S8, Figure S3). Greater vaccination administration naturally led to greater numbers of averted cases, deaths, and DALYs, but the order of impact of the different strategies remained the same.

To assess the impact of targeting the oldest age groups for vaccination, we evaluated vaccination in persons aged ≥60 years who were not healthcare workers or residing in a long-term care facility (Table 2). This population included 7.2 million individuals, and vaccination was estimated to avert approximately 80% of counterfactual deaths and 60% of counterfactual DALYs over 6 months.

**Table 2.** Simulated impact of targeting COVID-19 vaccinations by age.

| **Age (years)** | **Population vaccinated, N (%)** | **Cases averted, N (%)** | **Deaths averted, N (%)** | **DALYs averted, N (%)** |
| --- | --- | --- | --- | --- |
| 80+ | 1290094 (3) | 8300 (2) | 3100 (34) | 20000 (13) |
| 70-79 | 2200746 (6) | 19000 (5) | 2100 (24) | 29000 (19) |
| 60-69 | 3671923 (10) | 49000 (12) | 1900 (21) | 39000 (26) |
| 50-59 | 4614529 (12) | 65000 (16) | 1100 (12) | 32000 (21) |
| 40-49 | 4832598 (13) | 62000 (15) | 480 (5) | 18000 (12) |
| 30-39 | 5068999 (14) | 67000 (16) | 210 (2) | 9900 (6) |
| 20-29 | 5539817 (15) | 66000 (16) | 79 (1) | 4600 (3) |

\* Excludes residents of long-term care facilities (skilled nursing facilities and assisted-living facilities).

### Targeting vaccine allocation by multiple risk factors

To assess the potential benefit of targeting vaccinations by more than one risk factor, we simulated the impact of targeting by both age and county, and by both age and special population. Targeting by both age and special population performed equivalently to targeting purely by age in terms of the proportion of overall DALYs averted (both 38%, 95% UI 37-39%), but targeting by both age and county averted a higher proportion of DALYs (46%, 95% UI 45-47%). We also estimated the optimal vaccine allocation (in terms of DALYs averted) based on the combination of all modelled risk factors (demographic, special population status, and comorbidity status) (Figure 4). We identified that older individuals would still be prioritized under optimal allocation, but the proportion of individuals vaccinated would also vary considerably by county of residence, sex, race/ethnicity, and special population and comorbidity status. In particular, risk of DALYs was higher for residents of certain counties (including Imperial County, San Joaquin County, Stanislaus County, Los Angeles County, and Tulare County), Hispanic/Latinx and non-Hispanic Black individuals, certain special populations (prisoners and persons experiencing homelessness), individuals with comorbidities, and males. In some cases, higher risk of DALYs with older age was offset by higher risk on another factor, such as county of residence or race/ethnicity. For example, our estimates suggest vaccinating a higher proportion of 50-to-59-year-old residents of Imperial County than ≥80-year-old residents of San Francisco County (Figure 4A), and a higher proportion of 60-to-69-year-old Hispanic/Latinx individuals than ≥80-year-old non-Hispanic White individuals to avert the most DALYs (Figure 4C). Allocating vaccinations optimally according to all risk factors, i.e., prioritizing individuals in intersections of higher risk groups such as older Hispanic/Latinx individuals in the highest-risk counties, resulted in a significantly higher proportion of DALYs being averted (53%, 95% UI 52-54%) than any of the strategies that prioritized by a single risk factor (24-38%).

**Figure 4.**
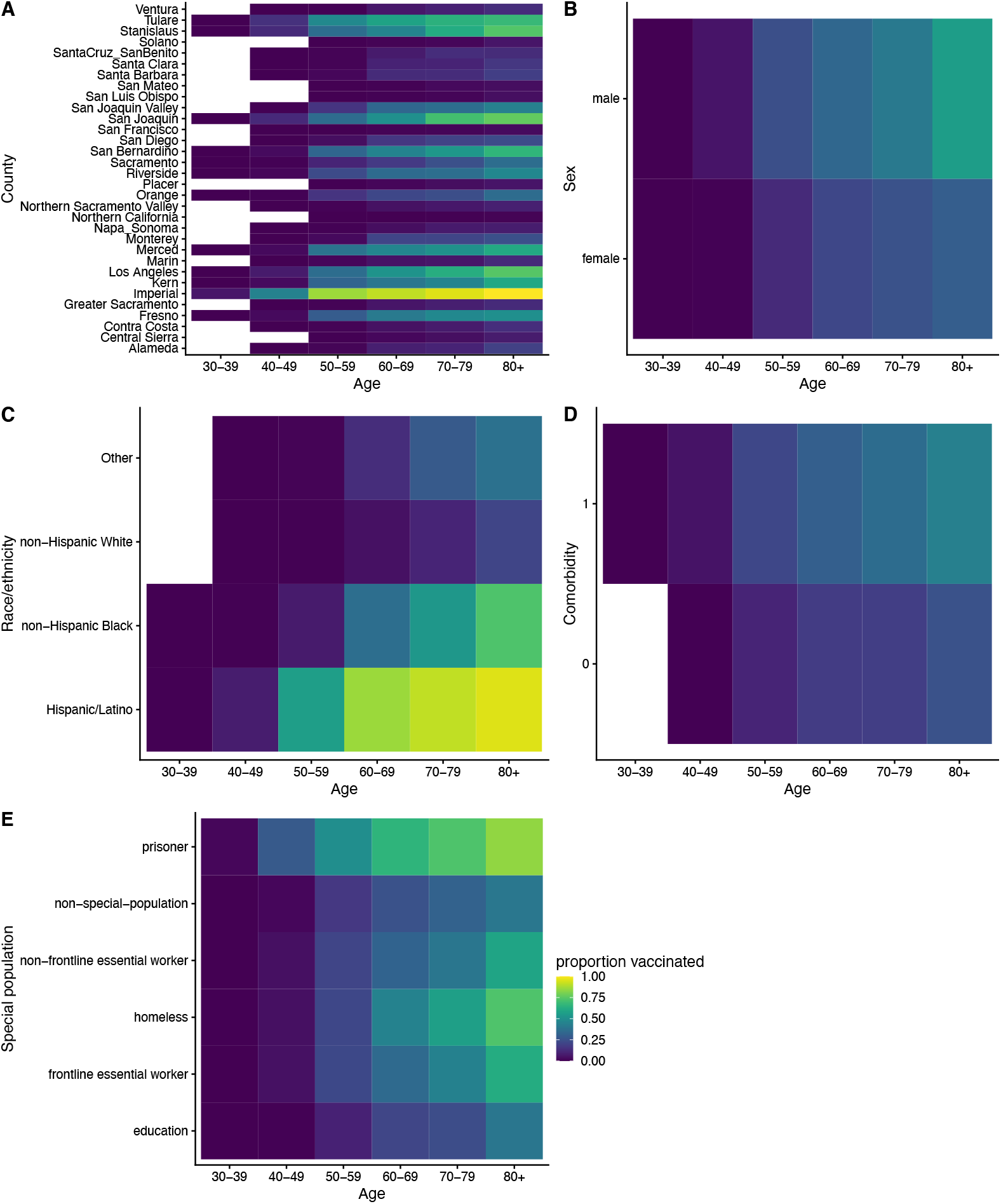
Optimal vaccine allocation under simultaneous targeting by multiple risk factors. Plots show the proportion of individuals in each (A) age-and-county group, (B) age-and-sex group, (C) age- and-race/ethnicity group, (D) age-and-comorbidity-status group, and (E) age-and-special population group vaccinated under optimal distribution of the first 5 million vaccinations in California. Lighter yellow colors show where higher proportions of individuals are vaccinated, darker blue colors where lower proportions of individuals are vaccinated. We assume all healthcare workers, skilled nursing facility residents and assisted living facility residents are vaccinated first before essential workers and other groups as per the US CDC recommendation. Vaccinations are optimally allocated to avert DALYs across all risk factors (demographic, special population, and comorbidities). Comorbidity status is treated as binary (0 = no comorbidity, 1 = any comorbidity). The proportions of individuals vaccinated under optimal vaccine distribution for each of the individual risk factors are shown in Figure S5.

### Sensitivity analyses

We varied the vaccine efficacy profile to determine its effect on the impact of the different vaccination strategies (Figure S6). With a vaccine with 60% efficacy against death instead of 95% efficacy, all prioritization strategies had lower impact (only 15-23% of DALYs were averted under single-risk-factor strategies compared to 24-38%), but their order of impact was the same for all vaccine availabilities. With a vaccine with 20% efficacy, all prioritization strategies had lower impact still (only 5-7% of DALYs were averted) and the relative differences between strategies were smaller. The order of impact of the strategies was the same and their relative impact very similar when vaccinations were prioritized according to risk of QALY loss rather than risk of DALY burden (cf. Figure S7 and Figure 3, and Table S9 and Table 1).

## Discussion

In this study, we have developed an approach for assessing the benefit of targeting allocation of limited initial vaccine doses during a pandemic by multiple risk factors. We have used the framework to compare the impact on key health outcomes of different prioritization strategies for early COVID-19 vaccine distribution, using California as a case example. By considering variation in risk of infection and death according to multiple risk factors (including age, location, sex, occupation, race/ethnicity and comorbidities) our study stands in contrast to many other analyses of COVID-19 vaccine prioritization ^16–22^, which have tended to focus on prioritization by a single risk factor. In agreement with other prioritization analyses ^16–22^, we find that, among strategies targeting by a single risk factor, prioritizing older individuals for vaccination minimizes deaths and DALYs when vaccine supply is limited. However, we find that targeting early vaccine distribution simultaneously by age and one or more other risk factors, such as location, has the potential to avert significantly more DALYs from COVID-19 than targeting vaccination purely by age. In an ongoing epidemic/pandemic, the consequences of prioritization of available vaccine doses are critical differences in mortality and morbidity, especially from the point of view of maintaining functioning of the health system. This is still the case for the COVID-19 pandemic, since vaccination coverage in some low-income countries is still very low and new variants with higher transmissibility and immune escape properties have emerged that pose a considerable threat to these countries.

Here we focused on identifying which groups should be prioritized for vaccination after initial allocation to healthcare workers and long-term care facility residents ^12^, given the high healthcare burden of COVID-19 and high IFR among older individuals. We found that to minimize DALYs and deaths due to COVID-19, older individuals (≥60 years) and those with comorbidities should be targeted for vaccination before essential workers and other special populations.

Our findings for COVID-19 are largely driven by the strong age-dependence of the COVID-19 infection fatality rate (Table S10), as well as differential risk by special population and occupation (Table S4). The age distribution of cases is skewed towards younger ages, with a peak in case numbers in 20-to-29-year-olds in the US, while the age distribution of deaths is strongly skewed towards older individuals, with the highest death rates among individuals ≥80 years of age (Figure 1C-D). As a result, there is a trade-off between minimizing cases (and therefore transmission) and minimizing deaths under different prioritization strategies. Targeting older individuals averts more deaths but fewer cases as it captures a larger proportion of those with high death risk. In contrast, strategies that target essential workers and special populations, who (excluding long-term care facility residents) tend to be younger but at higher risk of infection, avert more cases but fewer deaths. Patterns for DALYs averted are similar to those for deaths averted, since our estimates of DALY burden are driven mainly by mortality and not morbidity. We note, however, that we have not included morbidity from long-term sequelae of COVID-19 in our DALY estimates due to lack of available data ^35^, and that this may introduce some bias in our results towards greater impact in older individuals.

Our framework could be applied to vaccine prioritization in low-income countries that currently have low vaccine coverage. Data on demographic structure and risk factors would need to be updated, and this may affect the results to some extent, but age is likely to remain the primary risk factor for targeting vaccination given the consistency and strength of the age dependence in the infection fatality rate across countries.

Our results are in broad agreement with the findings of other modelling studies of vaccine prioritization ^16–22^ and the NASEM allocation framework ^10^ and the CDC ACIP prioritization recommendations ^13^, despite differences in analysis approaches. Most other modelling studies have used age-structured population-level COVID-19 transmission dynamic models to assess the impact of different age- and occupation-based targeting strategies ^16–21^, and have also concluded that deaths and years of life lost are minimized by targeting older individuals (aged over 60), while cumulative infection incidence is minimized by targeting younger individuals and workers at higher risk of infection, such as essential workers. The strength of our analysis is the data-driven approach we use to incorporate detailed heterogeneity in COVID-19 risk by demographic factors (age, location, sex, occupation, and race/ethnicity) and comorbidities. Our analysis suggests that targeting vaccinations purely by age averts a significant proportion of COVID-19 deaths and DALYs over the first 6 months of vaccine rollout (in our simulations for California it averted 40% of DALYs with sufficient doses to vaccinate 13% of the population), but that further targeting of vaccinations by location, sex, race/ethnicity, special population status, and comorbidity status would provide additional benefit (averting over half of the DALY burden with 13% vaccination coverage). However, the benefits of such a strategy would have to be weighed against increased logistical complexity and potentially cost of implementation before it could be adopted.

This analysis has limitations that should be considered. We used a static model to assess vaccination impact, which only accounts for direct effects of vaccination (protection from infection and severe disease), and not indirect effects through reduction in transmission. We took this approach in order to be able to include a detailed stratification of COVID-19 death risk and as our analysis focuses on allocation of the first available vaccinations (with relatively low coverage in the population), for which reduction in transmission may be relatively limited. Given evidence that vaccines are effective at reducing transmission and the emergence of more transmissible SARS-CoV-2 variants, consideration needs to be given to switching to vaccinate high-transmission groups, i.e., younger individuals and essential workers ^17,18^. We used the average baseline hazard rate for COVID-19 death from data up to the end of 2020 to simulate numbers of deaths, cases and infections over the first 6 months of 2021, which underestimated incidence in California due to increases in cases and deaths at the end of 2020. However, the focus of this study was not forecasting of COVID-19 cases and deaths, but comparison of the relative impact of different vaccination strategies, which is not affected by the baseline death rate in our model. We modelled vaccine protection as occurring immediately rather than in a phased rollout. Although this will have affected the precise quantitative estimates of clinical cases, deaths and DALYs averted under the different prioritization strategies, it will not have affected the ranking of the different prioritization strategies by impact. We have not compared the cost-effectiveness of the different prioritization strategies considered, despite potential differences in costs due to variability in difficulty of vaccine delivery. However, targeting by age is logistically more straightforward than targeting essential workers or non-healthcare-worker non-long-term-care-facility special populations, since existing programs and mechanisms for delivering age-targeted vaccination can be leveraged ^25^. We included assumptions to estimate infections and clinical cases which may introduce bias based on data used to calibrate the infection fatality rate for California, since there is likely under-reporting of clinical cases in the available case data and relatively limited seroprevalence data available for validation. Nonetheless, as the estimated contribution of infections and clinical cases to the overall DALY burden is small compared to deaths, this is unlikely to have affected the overall ranking of the different prioritization strategies. Our analysis only considered health outcomes and not equity measures, but there are considerable inequities in COVID-19 burden by race, ethnicity, and socioeconomic status that should also be assessed when prioritizing vaccinations ^36–38^.

Our detailed analysis of risk of mortality and morbidity from COVID-19 and estimation of impact of different vaccination prioritization strategies highlights the strength of age-based targeting as a strategy for averting COVID-19 deaths and the potential benefit of targeting vaccinations by other risk factors in addition to age. Given many low-income countries still have low vaccination coverage, ensuring that vaccines reach those most at risk of poor health outcomes from COVID-19 should remain a focus of the global vaccine rollout. Where vaccine supply is limited and logistical challenges in vaccine delivery persist, age-based targeting offers a viable means of achieving this goal. If operational challenges can be overcome, more granular vaccination strategies that overlap age with other risk factors can be adopted.

Decisions on who should receive the first available vaccinations in a country during an epidemic are complex. The choice of the primary outcome of importance for vaccine prioritization must ultimately be determined by stakeholders, weighing together all available evidence and the priorities of their health system. Nevertheless, the framework provided by this study can be used to support this decision-making process since it enables comparison of the impact of different prioritization strategies, including those that involve multiple risk factors.

## Data Availability

All analysis code is available online at https://github.com/LloydChapman/COVIDVaccineModelling. The California Department of Public Health case data required for fitting the Poisson regression model contain personally identifiable information and therefore cannot be made freely available. Individuals interested in accessing the data should contact the California Department of Public Health. The simulated population data required to run the vaccine prioritization simulations is available at http://doi.org/10.5281/zenodo.4516526.

## Acknowledgements

We are grateful to the California Department of Public Health for sharing the data used in this article, and sincerely thank all individuals involved in collecting and curating the data and those who helped with data access and interpretation, in particular Dr Jason Vargo and Dr William Wheeler. We thank Rhett Hilary, Svetlana Milter and Max Murphy at the University of California, San Francisco for IT support and Marinella Capriati for helpful discussions. We would also like to thank all those involved in the ongoing response to the COVID-19 pandemic in California.

## Author contributions

NCL, KBD, GRW and RS conceived the study. LACC, PS and NCL led the data collection activities with support from PBS and TML. LACC performed the data analysis with support from PS and NCL. LACC, PS and NCL drafted the manuscript. PBS, TML, KBD, GWR, IRB, RS contributed to interpretation of the results and edited the manuscript. All authors read and approved the final manuscript.

## Funding

This study was supported by funding from the California Department of Public Health. NCL is supported by the University of California, San Francisco (Department of Medicine). The funding organizations had no role in the design of the study; analysis and interpretation of the data; and preparation of the manuscript; or the decision to submit the manuscript for publication. This work represents the viewpoints of the authors alone and not necessarily those of the California Department of Public Health.

## Consent for publication

Not applicable.

## Competing interests

The authors declare that they have no competing interests or disclosures. NCL reports personal fees from the World Health Organization for unrelated work.

## Notes

### Competing Interest Statement

The authors have declared no competing interest.

### Author Declarations

The study was approved by the University of California, San Francisco Institutional Review Board.

### Summary of Updates

We now account for different vaccine efficacies against infection and death, waning of vaccine-induced immunity over time since infection, and the possibility of reinfection in the simulations of the different vaccine prioritization strategies.

