## Supplementary Information for "Risk factor targeting for vaccine prioritization during the COVID-19 pandemic"

### Supplementary methods

*Simulated California population*

We simulated the entire population of California (N=39,148,760) with information for each individual on:

- demographic characteristics: age, sex, race, ethnicity and county of residence;
- unique special population status: healthcare worker (HCW), incarcerated individual (state/federal/local/youth prisoner), skilled nursing facility (SNF) resident, assisted living facility (ALF) resident, education worker, person experiencing homelessness, frontline essential worker, non-frontline essential worker;
- binary status on each of seven comorbidities: asthma, diabetes, heart disease, heart failure, hypertension, obesity, active tobacco use.

Age-, sex- and race-specific population totals for each of the 58 counties in California from the 2018 American Community Survey (ACS) 5-year estimates (1) were used to assign age category (0-4, 5-9, 10-14, 15-17, 18-19, 20-24, 25-29, 30-34, 35-44, 45-54, 55-64, 65-74, 75-84, 85+ years), sex and race for individuals of each county (see Table S1 for variable definitions). Integer ages within each age group were simulated by uniform random sampling. To incorporate binary ethnicity status $E$ (Hispanic/Latino, non-Hispanic/Latino), we estimated the number of individuals of each ethnicity in each county-age-race group (denoted by subscript $C, A, R$), $N_{C,A,R,E}$, as:

$$N_{C,A,R,E} = N_{C,A,R}\,p_{E|C,A}\,p_{E|C,R}$$

where $p_{E|C,A}$ is the estimated proportion of the age group in the county that is of ethnicity $E$ and $p_{E|C,R}$ is the proportion of the race group in the county that is of ethnicity $E$ based on the ACS data, and randomly assigned this number of individuals in the county-age-race group to be of ethnicity $E$.

Overall population estimates for each of the special populations and their sources are given in Table S2. Special population status was assigned by drawing from a categorical distribution with probabilities for membership of each of the special populations dependent on one or more demographic factors. These probabilities were derived as follows. The probabilities of being a HCW and of being an education worker in each county-age-sex group, $p_{HCW|C,A,S}$ and $p_{educ|C,A,S}$ (subscript $S$ denoting sex), were estimated as:

$$p_{X|C,A,S} = \frac{N_{X,C,S}\,p_{A|X}}{N_{C,A,S}}$$

where $X \in \{HCW, educ\}$, $N_{X,C,S}$ is the number of HCWs/education workers in each county-sex group according to the ACS data, $p_{A|X}$ is the proportion of HCWs/education workers who are of age group $A$ according to data from the Bureau of Labour Statistics (2), and $N_{C,A,S}$ is the number of individuals in the county-age-sex group. The probabilities of being a prisoner and of being a SNF resident in each age-sex-race-ethnicity group, $p_{prison|A,S,R,E}$ and $p_{SNF|A,S,R,E}$, were estimated as:

$$p_{Y|A,S,R,E} = \frac{N_{Y,A,S}p_{R|Y}p_{E|Y}}{N_{A,S}p_R p_E}$$

where $Y \in \{prison, SNF\}$, $N_{Y,A,S}$ is the state-level prisoner/SNF population of age $A$ and sex $S$, $N_{A,S}$ is the state-level population of age group $A$ and sex $S$, $p_{R|Y}$ and $p_{E|Y}$ are the state-level proportions of prisoners/SNF residents who are of race $R$ and ethnicity $E$ respectively, and $p_R$ and $p_E$ are the corresponding proportions for California residents.

The total ALF population was estimated as approximately 150,000 from the current capacity of ALFs in California (185,000) (3) and data on the occupancy of ALFs from the National Center for Health Statistics' (NCHS) National Study of Long-Term Care Providers (NSLTCP) 2015-16 (4), which suggests 81% occupancy of ALFs in California. The probability of being an ALF resident in age group $A$, $p_{ALF|A}$, was then estimated as:

$$p_{ALF|A} = \frac{N_{ALF}p_{A|ALF}}{N_A}$$

where $N_{ALF}$ is the total ALF population, $N_A$ is the population of age group $A$, and $p_{A|ALF}$ is the proportion of ALF residents who are in age group $A$, taken from the NCHS NSLTCP, assuming that all ALF residents are ≥55 years-of-age.

The probability of being a person experiencing homelessness in each county-age-sex-race group was calculated as:

$$p_{homeless|C,A,S,R} = \frac{N_{homeless,C,A}p_{S|homeless}p_{R|homeless}}{N_{C,A}p_S p_R}$$

where $N_{homeless,C,A}$ is the number of people experiencing homelessness in the county-age group from the US Department of Housing and Urban Development's (DHUD) Continuum of Care homelessness data (5), $N_{C,A}$ is the county-age group population, $p_{S|homeless}$ is the proportion of people experiencing homelessness in California who are of sex $S$ from the DHUD data, and $p_{R|homeless}$ is the proportion of the homeless population who are of race $R$ based on Los Angeles Almanac homelessness data (6).

The probabilities of being a frontline essential worker and of being a non-frontline essential worker in each age group were estimated as follows. The national-level percentages of workers in each age group in each of the 5 broad categories of occupations used by the US Bureau of Labor Statistics (Management and professional, Service, Sales and Office, Natural resources, Production and transportation) (2) who are frontline and non-frontline essential workers was calculated using the CDC classification of frontline and non-frontline essential workers (7). These percentages were then multiplied by the percentages of each age group in California who are employed according to data from the 2018 ACS.

We estimated the probability of an individual in each county-age-sex-race group having each of the comorbidities ($p_{comorb|C,A,S,R}$) using data from the California Health Interview Survey (CHIS) (8) on prevalence of each comorbidity in each county by age ($p_{comorb|C,A}$), and that at the state level by sex, by race, and overall ($p_{comorb|S}$, $p_{comorb|R}$, and $p_{comorb}$), as:

$$p_{comorb|C,A,S,R} = p_{comorb|C,A} \frac{p_{comorb|S}}{p_{comorb}} \frac{p_{comorb|R}}{p_{comorb}}.$$

We used this probability to simulate binary status for each comorbidity for each individual in each county-age-sex-race group via a Bernoulli random draw. Obesity was defined as a body mass index (BMI) ≥30, where BMI was simulated using CHIS data on the state-level mean BMI and standard deviation for each age group, adjusted by sex and race as in the above formula. Each individual's BMI was simulated via a random draw from a truncated normal distribution with the age-sex-and-race-adjusted mean and standard deviation, and truncation limits of 5 and 50.

*COVID-19 case and death data*

De-identified individual-level data on 2,215,972 confirmed COVID-19 cases (individuals who tested PCR positive for SARS-CoV-2) in California up to December 30, 2020, was provided by the California Department of Public Health (CDPH). Among these cases, 28,175 individuals died due to COVID-19. The data included information on county of residence, age category, sex, race/ethnicity, date of positive PCR test result, and, where applicable, date of death (see Table S3 for variable definitions). Complete demographic data was available for 1,578,988 (71%) cases and 27,580 (98%) individuals who died.

*Overview of COVID-19 simulation model*

We simulated COVID-19 deaths, clinical cases, infections and disability-adjusted life-years (DALYs) using the following steps, each of which is described below in further detail:

1. Simulate COVID-19 deaths using Poisson regression model fitted to COVID-19 death data with death hazard rate adjusted by special population status and comorbidity status, and calculate associated DALYs.
2. Estimate infections from predicted deaths using published age- and sex-dependent COVID-19 infection fatality rate (IFR).
3. Estimate clinical cases from predicted infections using published age-dependent clinical fraction for COVID-19.
4. Calculate DALYs associated with infections and clinical cases.

**Table S1. Definitions and sources of variables in simulated California population**

| Variable | Values | Source |
|---|---|---|
| County of residence | Alameda, Alpine, Amador. Butte, Calaveras, Colusa, Contra Costa, Del Norte, El Dorado, Fresno, Glenn, Humboldt, Imperial, Inyo, Kern, Kings, Lake, Lassen, Los Angeles, Madera, Marin, Mariposa, Mendocino, Merced, Modoc, Mono, Monterey, Napa, Nevada, Orange, Placer, Plumas, Riverside, Sacramento, San Benito, San Bernardino, San Diego, San Francisco, San Joaquin, San Luis Obispo, San Mateo, Santa Barbara, Santa Clara, Santa Cruz, Shasta, Sierra, Siskiyou, Solano, Sonoma, Stanislaus, Sutter, Tehama, Trinity, Tulare, Tuolumne, Ventura, Yolo, Yuba | (1) |
| Age | Integer between 0 and 100 | (1) |
| Sex | Male, Female | (1) |
| Race | White, African American, American Indian or Alaska Native, Asian Alone, Native Hawaiian or Pacific Islander, Some other race alone, Two or more races | (1) |
| Ethnicity | Hispanic/Latino, non-Hispanic/Latino | (1) |
| Special population | Healthcare worker, Prisoner, Skilled Nursing Facility Resident, Assisted Living Facility Resident, Education Worker, Person Experiencing Homelessness, Frontline Essential Worker, Non-Frontline Essential Worker | See Table S2 |
| Asthma | 0, 1 | "AB40" variable in (8) |
| Diabetes | 0, 1 | "DIABETES" variable in (8) |
| Heart Disease | 0, 1 | "AB34" variable in (8) |
| Heart Failure | 0, 1 | "AB52" variable in (8) |
| Hypertension | 0, 1 | "AB29" variable in (8) |
| Obesity | 0, 1 | BMI ≥ 30 where BMI simulated using "BMI" variable in (8) (see text) |
| Active tobacco use | 0, 1 | "SMKCUR" variable in (8) |

**Table S2. Population estimates for special populations in California**

| Special population | Population estimate* | Source |
|---|---|---|
| Healthcare workers | 1,559,001 | (1) |
| Prisoners | 227,428 | (1) |
| Skilled nursing facility residents | 136,282 | (1) |
| Assisted living facility residents | 150,340 | (3,4) |
| Education workers | 962,684 | (1) |
| Persons experiencing homelessness | 132,427 | (5) |
| Frontline essential workers | 2,521,235 | (1,2,7) |
| Non-frontline essential workers | 6,994,107 | (1,2,7) |

* Note that population totals reflect values simulated as described in the text, which may not exactly match totals given in sources.

**Table S3. Definitions of variables in COVID-19 case and death data used in analysis**

| Variable | Definition | Values | Modifications |
|---|---|---|---|
| county_res | County of residence of case | Alameda, Central Sierra, Contra Costa, Fresno, Greater Sacramento, Imperial, Kern, Los Angeles, Marin, Merced, Monterey, Napa_Sonoma, Northern California, Northern Sacramento Valley, Orange, Placer, Riverside, Sacramento, San Bernardino, San Diego, San Francisco, San Joaquin, San Joaquin Valley, San Luis Obispo, San Mateo, Santa Barbara, Santa Clara, SantaCruz_SanBenito, Solano, Stanislaus, Tulare, Ventura | Counties with population <250,000 combined into single region (except Imperial), San Benito combined with Santa Cruz, Napa combined with Sonoma |
| age_cat | Age category | <10, 10-19, 20-29, 30-39, 40-49, 50-59, 60-69, 70-79, 80+ | |
| sex | Sex | Male, Female | |
| race_ethnicity | Combined race/ethnicity variable | Hispanic/Latino, non-Hispanic Black, non-Hispanic White, Other | |
| first_report_date | Date of first report | 02/03/2020 - 12/30/2020 | Randomly moved forward or back 1 day |
| date_of_death | Date of death | 02/05/2020 - 12/30/2020 | Randomly moved forward or back 1 day |

*Calibration of infection fatality rate for California*

To account for variation in the IFR across different settings, we recalibrated the age- and sex-dependent ensemble IFR estimate from (9) for California, using a combination of data on confirmed cases and deaths from CDPH up to late October, 2020; seroprevalence estimates for California up to October 8-13, 2020 (from 6 biweekly surveys of approximately 980 individuals), from the CDC's database of nationwide commercial laboratory seroprevalence surveys (10); and age-stratified data on numbers of COVID-19 deaths in California among long-term care facility (LTCF) residents (SNF and ALF residents) and non-LTCF residents provided by CDPH. The IFR estimate from (9) is for non-LTCF residents and LTCF residents account for approximately 30% of COVID-19 deaths in California, so we used the age-and-LTCF-stratified death data to estimate the number of deaths in each demographic risk group, of 19,501 cumulative deaths up to October 31, 2020, that were among non-LTCF residents. We then divided this number by the IFR to estimate the number of infections among non-LTCF residents in each demographic risk group, and multiplied by the estimated age-dependent clinical case fraction from (11) (which fitted an age-structured transmission model to age-stratified clinical case data from 6 countries to the estimate the clinical fraction) to estimate the number of clinical cases among non-LTCF residents. We calculated an overall multiplier for the IFR of 2.64 to match the cumulative number of clinical cases among non-LTCF residents from our model with the cumulative number of non-LTCF confirmed cases in the CDPH case line list up to October 22, 2020 (827,867) (accounting for a mean reporting-to-death time of 9 days calculated from the data). We note that this does not account for the fact that some confirmed cases up to October 22, 2020, were asymptomatic and that symptomatic cases were under-reported early in the pandemic. However, these two biases will have acted in opposite directions and information on whether confirmed cases were asymptomatic was unavailable, so we assume that the cumulative number of confirmed cases up to October 22, 2020, provides a reasonable approximation of the cumulative number of clinical cases up to that date. We verified that this approach gave a reasonable match between the age distribution of clinical cases from the model and that of confirmed cases (Figure 2A in main text), and between the age distribution of cumulative infections estimated from the model (2,433,000) and that estimated from the mean seroprevalence estimates for California up to October 8-13, 2020 (1,927,000) (using an approximately 12-day infection-to-reporting delay between the seroprevalence data centred on October 10 and case data calculated from the median onset-to-reporting delay in the data and incubation period (12)) (Figure 2B in main text). We note that the estimate of the number of individuals infected based on the seroprevalence data is likely to be an underestimate of the true number as it does not account for waning of antibodies (13).

As a further check on this approach, we performed a Poisson regression of cumulative case numbers against demographic risk factors (age, sex, race/ethnicity and county of residence) for both the estimated clinical case counts and confirmed case counts and compared the resulting parameter estimates. Overall, the two sets of parameter estimates agreed closely with each other (Figure S1). The hazard ratio estimates from the estimated clinical case counts were higher than those from the confirmed case counts for certain counties, possibly suggesting greater under-reporting of cases in these counties and/or higher IFRs, e.g., due to overburdened hospitals.

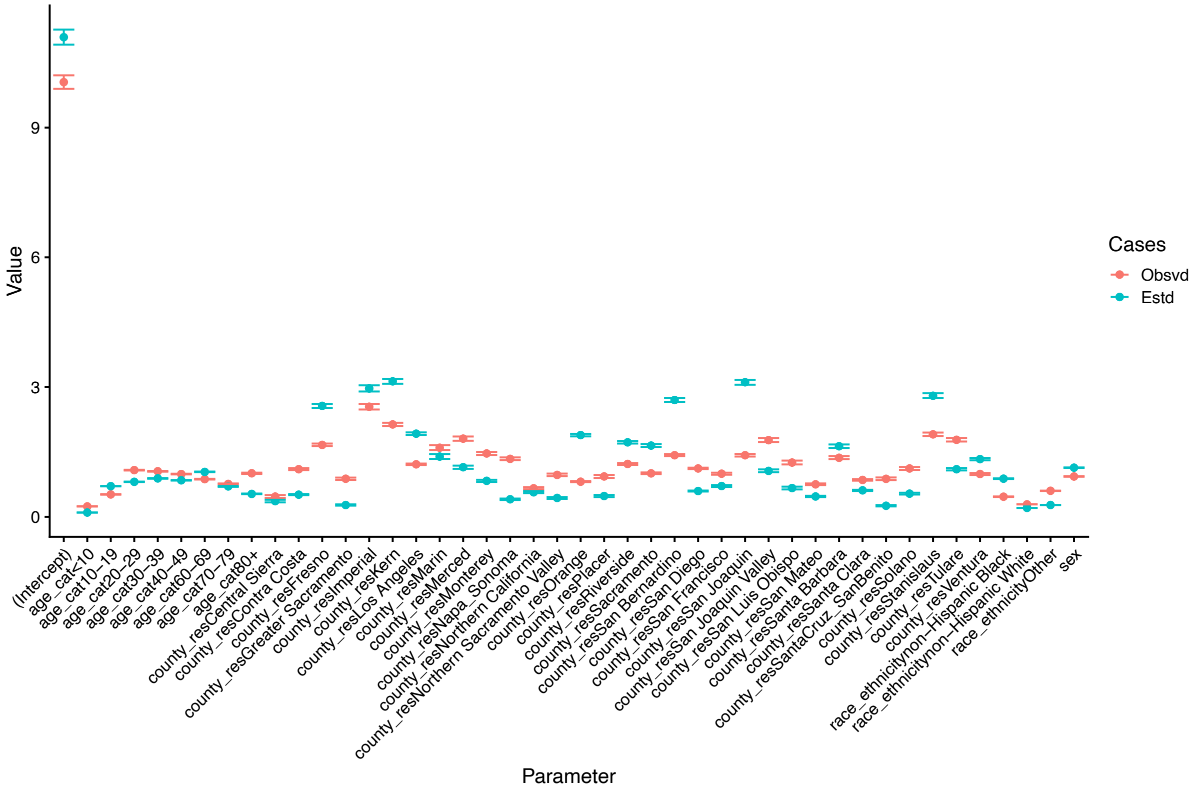

**Figure S1. Model calibration check.** Parameter estimates for Poisson regressions of COVID-19 case risk associated with different demographic risk factors for confirmed cases up to October 22, 2020, from the California Department of Public Health (red) and clinical cases estimated from our model (turquoise). Logarithm of total exposure time used as an offset variable, such that intercept represents COVID-19 case incidence rate (cases/100,000 person-days) among 50-59-year-old Hispanic/Latino females in Alameda County. Dots show maximum likelihood parameter estimates and error bars show 95% CIs.

*Poisson regression model for COVID-19 death risk*
We applied a Poisson regression model to the CDPH COVID-19 death data to estimate the hazard ratios for COVID-19 death associated with different demographic risk factors, namely county of residence, age, sex and race/ethnicity. The outcome variable was the cumulative number of deaths for each combination of the risk factors and the logarithm of the cumulative survival time for individuals with each combination of the risk factors was used as an offset variable to account for differences in time to death between individuals. Representing the different risk factors (county, age group, sex, race/ethnicity) by a vector of $p$ covariates $\mathbf{X} = (X_1, \dots, X_p)$ whose value is indexed by the index set $(i_1, \dots, i_p)$, and the total number of COVID-19 deaths and total survival time of individuals with covariate levels $(i_1, \dots, i_p)$ by $D_{i_1 \dots i_p}$ and $T_{i_1 \dots i_p}$ respectively (such that $D_{i_1 \dots i_p}/T_{i_1 \dots i_p}$ is the COVID-19 death rate among individuals with covariate levels $(i_1, \dots, i_p)$), the Poisson regression model is:

$$D_{i_1 \dots i_p} \sim \mathrm{Po}(\mu_{i_1 \dots i_p})$$

with

$$\log\,(\mu_{i_1 \dots i_p}) = \log\left(T_{i_1 \dots i_p}\right) + \log(\lambda_0) + \mathbf{X}^{\mathrm{T}}\boldsymbol{\beta},$$

where $\mu_{i_1 \dots i_p}$ is the conditional mean of the Poisson distribution for the cumulative number of deaths, $\boldsymbol{\beta} = (\beta_1, \dots, \beta_p)$ is a column vector of the covariate coefficients (such that $e^{\beta_i}$ is the hazard ratio for covariate $X_i$), and $\lambda_0$ is the baseline COVID-19 death rate in California (assumed constant). This model is equivalent to a proportional hazards model for time to COVID-19 death (14,15), with hazard rate:

$$\lambda_{i_1 \dots i_p} = \frac{\mu_{i_1 \dots i_p}}{T_{i_1 \dots i_p}} = \lambda_0 e^{\mathbf{X}^{\mathrm{T}}\boldsymbol{\beta}}$$

Hazard ratios and their standard errors were estimated using the `glm` function in `R` (version 4.0.3) (16). We compared parameter estimates for death data from different periods of time (the last 3 months and 6 months of 2020, and time from the first death to the end of 2020) to assess their stability over time.

*Adjustment of COVID-19 death risk by special population status and comorbidity status*
We adjusted risk of COVID-19 death by special population status and comorbidity status using published literature estimates for the relative risk of SARS-CoV-2 infection and death from COVID-19 for the special populations and hazard ratios for death given SARS-CoV-2 infection associated with the comorbidities (Table S4). For LTCF residents, we used estimates of age-specific relative risk of COVID-19 death calculated from the CDPH age-and-

LTCF-stratified data. For essential workers, we calculated estimates of the relative risk of COVID-19 death for frontline and non-frontline essential workers using estimates of relative risk of death by occupation from a recent analysis of excess mortality in California during the pandemic (17), categorising employment sectors according to the CDC classification of essential workers (7). For each special population $j$ for which data on relative risk of death was not available (healthcare workers, prisoners, education workers, and people experiencing homelessness), we derived an estimate of the relative risk of death, $RR_j$, by multiplying an estimate of relative risk of infection in the special population, $RR_{inf,j}$ (from published literature or calculated from publicly available data on cumulative incidence of confirmed cases in the special population), by an estimate of the relative risk of death given infection, $RR_{death|inf,j}$, derived from the age- and sex-specific IFR, to account for differences in age distributions between special populations:

$$RR_j = RR_{inf,j} RR_{death|inf,j} = RR_{inf,j} \frac{\overline{IFR_j}}{\overline{IFR}}$$

where $\overline{IFR_j}$ is the population-weighted average IFR for special population $j$ and $\overline{IFR}$ is the population-weighted average IFR for the general population. We assumed that comorbidities only affected risk of death once infected, and that they had a multiplicative effect on risk of death. The death rate for each demographic risk factor group $\lambda_{i_1 \dots i_p}$ was multiplied by the following factor to ensure the overall death risk remained the same (i.e. to account for the higher death risk for individuals in special populations and with comorbidities):

$$\frac{N_{i_1 \dots i_p}}{\sum_{j,k_1,\dots,k_7} RR_{jk_1 \dots k_7} N_{i_1 \dots i_p j k_1 \dots k_7}}$$

where $N_{i_1 \dots i_p}$ is the number of individuals with demographic risk factor levels $(i_1, \dots, i_p)$, $RR_{jk_1 \dots k_7}$ is the relative risk of death associated with special population $j$ (where $j = 0$ represents individuals that are not part of one of the 8 special populations) and comorbidity status $(k_1, \dots, k_7)$ (the particular combination of binary statuses for the 7 comorbidities), and $N_{i_1 \dots i_p j k_1 \dots k_7}$ is the number of individuals in each demographic-special-population-comorbidity-status subgroup.

*Estimation of number of individuals already infected*

We estimated the number of individuals already infected in each demographic risk group by dividing the observed cumulative number of deaths up to December 30, 2020, in each group by the age- and sex-specific recalibrated IFR for that group (see *Calibration of infection fatality rate for California*). For LTCF residents, we first multiplied the IFR by a frailty index representing increased risk of death given infection among LTCF residents compared to the general population, which we assumed to be 3 based on previous estimates (9,18). The probability of past infection in each age-sex-special-population group was calculated by dividing the estimated number of infected individuals by the group population size, and used to simulate whether or not individuals in the simulated population had been previously infected.

**Table S4. Estimates of relative risks of SARS-CoV-2 infection and COVID-19 death for special populations and hazard ratios for death given infection for different comorbidities**

| Parameter | Distribution/Value* | Source |
|---|---|---|
| Relative risk of SARS-CoV-2 infection for special population (Ref. = general population)** | | |
| Healthcare worker | $TN(3.4,0.04,3,4)$ | (19) |
| Prisoner | $TN(5.5,0.07,5,6)$ | (20) |
| Person experiencing homelessness | $TN(1.65,0.006,1.5,1.8)$ | (21,22) |
| Education worker | $TN(1.8,0.09,1.2,2.8)$ | (23) |
| Relative risk of COVID-19 death for special population (Ref. = general population) | | |
| Skilled nursing facility resident | Age-dependent | California Department of Public Health |
| Assisted living facility resident | Age-dependent | California Department of Public Health |
| Frontline essential worker*** | $TN(1.2,0.01,1,1.4)$ | (17) |
| Non-frontline essential worker*** | $TN(1.1,0.003,1,1.2)$ | (17) |
| Hazard ratio for COVID-19 death given infection for individual with comorbidity | | |
| Asthma | 1 | (24,25) |
| Diabetes | 1.51 | (26) |
| Heart disease | 1.16 | (24–26) |
| Heart failure | 1.77 | (25) |
| Hypertension | 1 | (25,26) |
| Obesity | 1.33 | (24) |
| Smoker | 1 | (24–26) |

* $TN(\mu,\sigma^2,a,b)$ = truncated normal distribution with mean $\mu$, non-truncated variance $\sigma^2$ and left and right truncation limits $a$ and $b$. Uncertainty bounds for relative risks of infection reflect a combination of statistical uncertainty (95% CIs for published estimates) and uncertainty due to variation in values from different sources. Death hazard ratios with 95% CI overlapping 1 in original source taken to be 1.

** Further details of the sources and calculations used to derive these values are given in File S1.

*** Estimated as mean of relative risks of death for workers in different employment sectors from (17) categorised into frontline vs non-frontline essential work using the CDC classification (7).

*Simulation of COVID-19 deaths*

To simulate COVID-19 deaths, we first calculated the cumulative probability of death for individuals in the simulated population over 6 months according to their demographic characteristics, special population status, comorbidities, simulated history of infection, and whether or not they were vaccinated. The cumulative probability of death over 6 months ($T = 182$ days) for a susceptible unvaccinated individual with covariate levels $(i_1,\dots,i_p)$ in special population $j$ with comorbidity status $(k_1,\dots,k_7)$ was:

$$p_{i_1\dots i_p j k_1\dots k_7} = 1 - e^{-\lambda_{i_1\dots i_p} RR_{jk_1\dots k_7} T}.$$

We assumed that the vaccine is 'leaky' and has efficacy $v_e(t) \in [0,1]$ against death that depends on time since vaccination $t$, i.e. reduces the relative risk of death at time $t$ after vaccination by a factor $(1 - v_e(t))$. We accounted for waning of vaccine-induced immunity against death by assuming that vaccine efficacy decreases exponentially at a rate $r$ with time since vaccination, i.e. $v_e(t) = v_{e0}e^{-rt}$, where $v_{e0} = 0.95$ is the initial vaccine efficacy. Based on an estimated reduction in efficacy of the Pfizer vaccine against death from a maximum of 98.2% 2-9 weeks post vaccination to 90.4% 20+ weeks post-vaccination (27), we used a value of $r = -\frac{1}{182}\log\left(1 - \frac{98.2-90.4}{98.2}\right) = 0.00045\ \text{day}^{-1}$. The cumulative probability of death over 6 months for a vaccinated individual not previously infected was therefore:

$$\begin{aligned} p_{i_1 \ldots i_p j k_1 \ldots k_7} &= 1 - e^{-\int_0^T (1-v_e(t))\lambda_{i_1 \ldots i_p} RR_{jk_1 \ldots k_7} dt} \\ &= 1 - e^{-\lambda_{i_1 \ldots i_p} RR_{jk_1 \ldots k_7}\left(T - \frac{v_{e0}}{r}(1-e^{-rT})\right)} \end{aligned}$$

Since a number of studies have shown that immunity acquired from previous infection wanes over time (28–31), we also accounted for potential reinfection and subsequent death amongst previously infected ($i$) individuals by modeling their cumulative probability of death over 6 months as:

$$p_{i_1 \ldots i_p j k_1 \ldots k_7, i} = 1 - e^{-\lambda_{i_1 \ldots i_p} RR_{jk_1 \ldots k_7}\left(T - \frac{i_{e0}}{r}(1-e^{-rT})\right)}$$

where $i_{e0}$ is the initial protection against death afforded by previous infection. In other words, we assumed that previous infection offers protection against death that wanes at the same rate as protection from vaccination and made the simplifying assumption that immunity from previous infection only began to appreciably wane at the start of 2021. We use $i_{e0} = v_{e0} = 0.95$, based on evidence of similar levels of protection against symptomatic infection from prior natural infection and two doses of a viral vector vaccine or mRNA vaccine (32).

The number of past infections, $I_{past,g}$, and future deaths amongst those not previously infected, $D_g$, and those previously infected, $D_{g,i}$, in each demographic-special-population-comorbidity risk group $g = (i_1, \ldots, i_p, j, k_1, \ldots, k_7)$ was simulated as

$$\begin{gathered} I_{past,g} \sim Bin(N_g, p_{past,g}), \\ D_g \sim Bin(N_g - I_{past,g}, p_g), \\ D_{g,i} \sim Bin(I_{past,g}, p_{g,i}), \end{gathered}$$

where $p_{past,g}$ is the probability of previous infection for individuals in the risk group (estimated as described above). We ran 1000 simulations of past infections and future deaths for the full California population ($N = 39{,}148{,}760$) to account for stochastic uncertainty. We also accounted for uncertainty in the estimated death rate and relative risks of infection for the special populations, by drawing values for $\log(\lambda_{i_1 \ldots i_p})$ from truncated normal distributions with bounds and standard deviations derived from the 95% confidence intervals (CIs) for the regression coefficients (Table S6), and values for the relative risks from truncated normal distributions with bounds and standard deviations derived from uncertainty in literature estimates (Table S4).

*Estimation of infections and clinical cases*
The cumulative number of SARS-CoV-2 infections in each demographic-special-population-comorbidity risk group was calculated from the simulated number of deaths by dividing by the recalibrated age- and sex-dependent IFR, adjusted for vaccinated individuals to account for different vaccine efficacies against infection and death:

$$I_g = \begin{cases} \dfrac{D_g}{IFR_g} & \text{for unvaccinated susceptible individuals} \\ \dfrac{D_g}{\dfrac{1-\overline{v_e}}{1-\overline{v_{ei}}} IFR_g} & \text{for vaccinated individuals} \end{cases}$$

where $\overline{v_e}$ and $\overline{v_{ei}}$ are the average vaccine efficacies against death and infection over the first 6 months of the rollout, and $IFR_g$ is the IFR for individuals in risk group $g$ given their age and sex. As for the vaccine efficacy against death, we assumed that the vaccine efficacy against infection decreases exponentially over time, $v_{ei}(t) = v_{ei0}\, e^{-r_i t}$, but from initial efficacy $v_{ei0}$ at rate $r_i = -\frac{1}{182}\log\left(1 - \frac{92.4-69.7}{92.4}\right) = 0.0015\ \text{day}^{-1}$ (based on an estimated decline in the efficacy of the Pfizer vaccine against symptomatic infection from 92.4% 1 week after vaccination to 69.7% 20+ weeks after vaccination (27)). The average vaccine efficacies are therefore given by:

$$\overline{v_e} = \frac{v_{e0}}{rT}\left(1 - e^{-rT}\right)$$
$$\overline{v_{ei}} = \frac{v_{ei0}}{r_i T}\left(1 - e^{-r_i T}\right)$$

We used $v_{ei0} = 0.85$ based on published estimates of two-dose efficacy of the Pfizer vaccine against infection (Table S7 in (33)). We assumed that levels of protection against infection and death from previous infection are the same as those from vaccination, such that the cumulative number of reinfections among previously infected individuals could be calculated as:

$$I_{g,i} = \frac{D_{g,i}}{\dfrac{1-\overline{v_e}}{1-\overline{v_{ei}}} IFR_g}.$$

Numbers of clinical cases were then calculated from estimated numbers of infections by multiplying by the estimated age-dependent clinical case fraction from (11), adjusted for vaccinated individuals to account for vaccine-induced protection against clinical symptoms given infection. In the sensitivity analyses for different vaccine efficacies (see *Methods* in main text), we assumed the same scaling of vaccine efficacy against infection to vaccine efficacy against death as in the main analysis, i.e. $v_{ei}/v_e = 0.85/0.95 = 0.89$.

*Calculation of DALYs*
DALYs associated with illness and/or death due to a disease are given by the sum of years of life lost (YLL) and years lived with disability (YLD):

$$DALYs = YLL + YLD$$

and thus provide a composite measure of mortality and morbidity. Years lived with disability depend on the duration of illness $d$ and the disability weight $w \in [0,1]$ associated with the illness (where 0 represents perfect health and 1 represents death):

$$YLD = wd.$$

Total DALYs associated with simulated COVID-19 deaths, $DALYs_d$, were calculated as:

$$DALYs_d = \sum_g D_g(\overline{L_g} + w_s d_s),$$

i.e. by adding the average remaining life expectancy $\overline{L_g}$ of individuals in risk group $g$ who died, calculated from 2018 US life tables (34) and the age, sex, and race/ethnicity distribution within the risk group, to YLD associated with severe illness from COVID-19, $w_s d_s$, multiplying by the number of deaths in the risk group, $D_g$, and summing over all risk groups. Total DALYs from clinical cases, $DALYs_c$, and subclinical infections, $DALYs_s$, were calculated as:

$$DALYs_c = I_c w_m d_m$$
$$DALYs_s = (I - I_c) w_{mi} d_{mi}$$

where $I_c$ and $I$ are the estimated numbers of clinical cases and overall infections respectively, and $w_m d_m$ and $w_{mi} d_{mi}$ are the YLD associated with clinical and subclinical infection respectively. We used estimates of disability weights for acute episodes of mild, moderate and severe illness from (35) for subclinical infection, clinical infection and death from COVID-19 respectively: $w_{mi} = 0.005, w_m = 0.053, w_s = 0.210$. For the corresponding durations of illness $d_{mi}$, $d_m$ and $d_s$, we used data on symptom duration stratified by symptom severity from a study of 273 COVID-19 outpatients in Atlanta, Georgia (36) and data from other studies and systematic reviews (37–39) that suggests median durations of symptoms for subclinical infection, clinical infection, and severe illness prior to death are 7 days, 10 days and 18 days respectively.

Approximate 95% uncertainty intervals (UIs) for predicted outcomes were calculated as the 2.5%-97.5% quantiles of the distributions of the simulated outcomes.

*Calculation of QALYs*

As a sensitivity analysis, we also considered prioritising vaccine allocation under each vaccination strategy (see *Methods* in main text) to maximize quality-adjusted life years (QALYs) saved instead of DALYs averted. Total QALY losses due to death, $QALYs_d$, were calculated as:

$$QALYs_d = \sum_g D_g E_g$$

where $E_g$ is the quality-adjusted value of the average remaining life expectancy, $\overline{L_g}$, of an individual in risk group $g$. The quality-adjusted life expectancy is given by:

$$E_g = \sum_{i=1}^{\overline{L_g}} Q_{\overline{a_g}+i}$$

where $Q_a$ is the quality of life weight for an individual of age $a$, and $\overline{a_g}$ is the average age of an individual in risk group $g$. We used values for the US from Table 3.6 of (40) for $Q_a$ (Table S5). We calculated total QALYs lost due to clinical and subclinical infections, $QALYs_c$ and $QALYs_s$, as:

$$QALYs_c = I_c u$$
$$QALYs_s = 0.07(I - I_c)u$$

where $u$ is the QALYs lost per clinical case, which we took to be 0.007 (based on values of 0.007 in (41) and 0.0075 in (42)), and the disutility of subclinical infection has been assumed to be approximately 7% of that of clinical infection, in line with its relative disability weight and duration in our DALY calculations.

**Table S5. Age-specific quality of life weights for US population derived from** (40) **and** (43)

| Age group (years) | Quality of life weight, $Q_a$ |
|---|---|
| 0-9 | $0.948$ |
| 10-19 | $0.948$ |
| 20-29 | $(0.924 + 0.912)/2 = 0.918$ |
| 30-39 | $(0.912 + 0.889)/2 = 0.906$ |
| 40-49 | $(0.889 + 0.855)/2 = 0.872$ |
| 50-59 | $(0.855 + 0.830)/2 = 0.843$ |
| 60-69 | $(0.830 + 0.817)/2 = 0.824$ |
| 70-79 | $(0.817 + 0.755)/2 = 0.786$ |
| 80+ | $0.755$ |

*Vaccine prioritization*

For all vaccine prioritization strategies, we assumed that all HCWs and LTCF residents were vaccinated first as per the CDC guidelines for the first phase of the vaccine rollout (44). For the remainder of the population, we calculated the expected average risk of DALYs per person per day for all subgroups under the prioritization strategy, and then ranked subgroups in descending order of DALY risk. Vaccines were then allocated to these subgroups in this order and randomly within each subgroup until the total number of vaccines available, $n_v$, was reached. So, e.g., for age targeting, after vaccination of HCWs and LTCF residents, vaccinations were allocated to the age group with the highest average DALY risk and then to age groups with progressively decreasing DALY risk until the vaccine quota was reached. The random allocation of vaccinations within subgroups was varied in each of the 1000 simulations to account for uncertainty from variation in the individuals vaccinated before the vaccine quota was met. Three different initial vaccine quotas were considered ($n_v = 2$ million, 5 million, 10 million), reflecting variation in initial vaccine availability.

Under special population targeting, essential workers (frontline and non-frontline) were grouped with the section of the population not belonging to any special population. Under essential worker targeting, frontline and non-frontline essential workers were grouped together and the rest of the non-HCW, non-LTCF-resident population was treated as one group, and allocation was assumed to be random within these two groups. For comorbidity targeting, we grouped individuals into those with any comorbidities and those with no comorbidities, and assumed allocation was random within these groups, on the basis that targeting by number of comorbidities would be practically and economically infeasible. Under age-and-special-population targeting, essential workers were included among the special populations targeted for vaccination. In all strategies, excess vaccines remaining after complete target group coverage were randomly allocated amongst the remaining population.

*Data and code availability*
All analysis code was developed in R version 4.0.3 (16) and is available online at https://github.com/LloydChapman/COVIDVaccineModelling. The CDPH case data required for fitting the Poisson regression model contain personally identifiable information and therefore cannot be made freely available. Individuals interested in accessing the data should contact CDPH. All the data required to run the vaccine prioritization simulations is available at http://doi.org/10.5281/zenodo.4516526.

**Supplementary results**
*Regression model parameter estimates*
Parameter estimates for the Poisson regression model (Table S6 and Figure S2) show significant variation in COVID-19 death risk across counties, with hazard ratios (HRs) relative to Alameda County varying from 0.38 for Northern California to 3.7 for Imperial County (when fitting to all data since the first recorded death on February 5, 2020). The hazard ratio estimates for age, sex and race/ethnicity reflect the increase in death risk with increasing age, higher death risk for males, and higher death risk for Hispanic/Latino individuals described in the main text. There was some variation in parameter estimates when fitting to different periods of past data (the last 3, 6, and 11 months of 2020), e.g., a few counties went from being lower risk than Alameda County to higher and vice versa, and there was a slight downward shift in the age distribution of deaths over time, but overall parameter estimates were highly consistent (Table S6).

*Vaccine availability*
The order of impact of the different prioritization strategies on DALYs, deaths and cases was robust to variation in the vaccine availability. However, with 2 million vaccinations instead of 5 million, i.e. only sufficient doses to vaccinate 5% of California's population rather than 13%, none of the prioritization strategies targeting by a single risk factor averted significantly more DALYs than random allocation over 6 months since the vast majority of the 2 million vaccinations were used up in vaccinating HCWs and LTCF residents (~1.85 million individuals in California) in the first phase of the rollout, and prioritization of the remaining 150,000 vaccinations made only a small difference to the numbers of DALYs averted. Prioritizing older individuals averted 18% (95% UI 18-19%) of the overall DALY burden under no vaccination, while prioritizing special populations, individuals with

comorbidities, essential workers and random allocation all averted 17-18% (Table S7 and Figure S3E). However, targeting across all risk factors simultaneously averted a higher proportion of DALYs (23%, 95% UI 22-23%). There was little difference in impact on cases between the different strategies – the percentage of cases averted was between 6% for all strategies (Table S7 and Figure S3A) – but allocating the vaccinations remaining after the first phase to older individuals averted more deaths (31%) than allocating them to special populations, individuals with comorbidities, or essential workers (all 28%).

With double the vaccine availability – 10 million vaccinations instead of 5 million, i.e., enough vaccine to vaccinate just over a quarter of California's population – age targeting averted by far and away the most DALYs of the strategies targeting by a single risk factor and 62% (95% UI 61-63%) of the simulated overall burden over 6 months (Table S8 and Figure S3F). This was principally due to averting a much higher proportion of deaths than any of the other strategies – 79% (95% UI 79-80%) compared to 54% (95% UI 53-56%) for comorbidity targeting as the next best performing strategy (Table S8 and Figure S3D). Special population targeting and essential worker targeting averted only 38% (95% UI 37-40%) and 35% (95% UI 34-36%) of the DALY burden respectively, due to only averting 46% (95% UI 44-47%) and 37% (95% UI 37-38%) of deaths. As for 5 million individuals vaccinated, age-and-county targeting and targeting simultaneously across all risk factors averted a higher proportion of DALYs (68% and 72% respectively) than any of the strategies targeting by a single risk factor. Of strategies targeting by a single risk factor, essential worker targeting averted the highest proportion of clinical cases (26%, 95% UI 22-29%), and age targeting averted the lowest proportion (22%, 95% UI 19-24%).

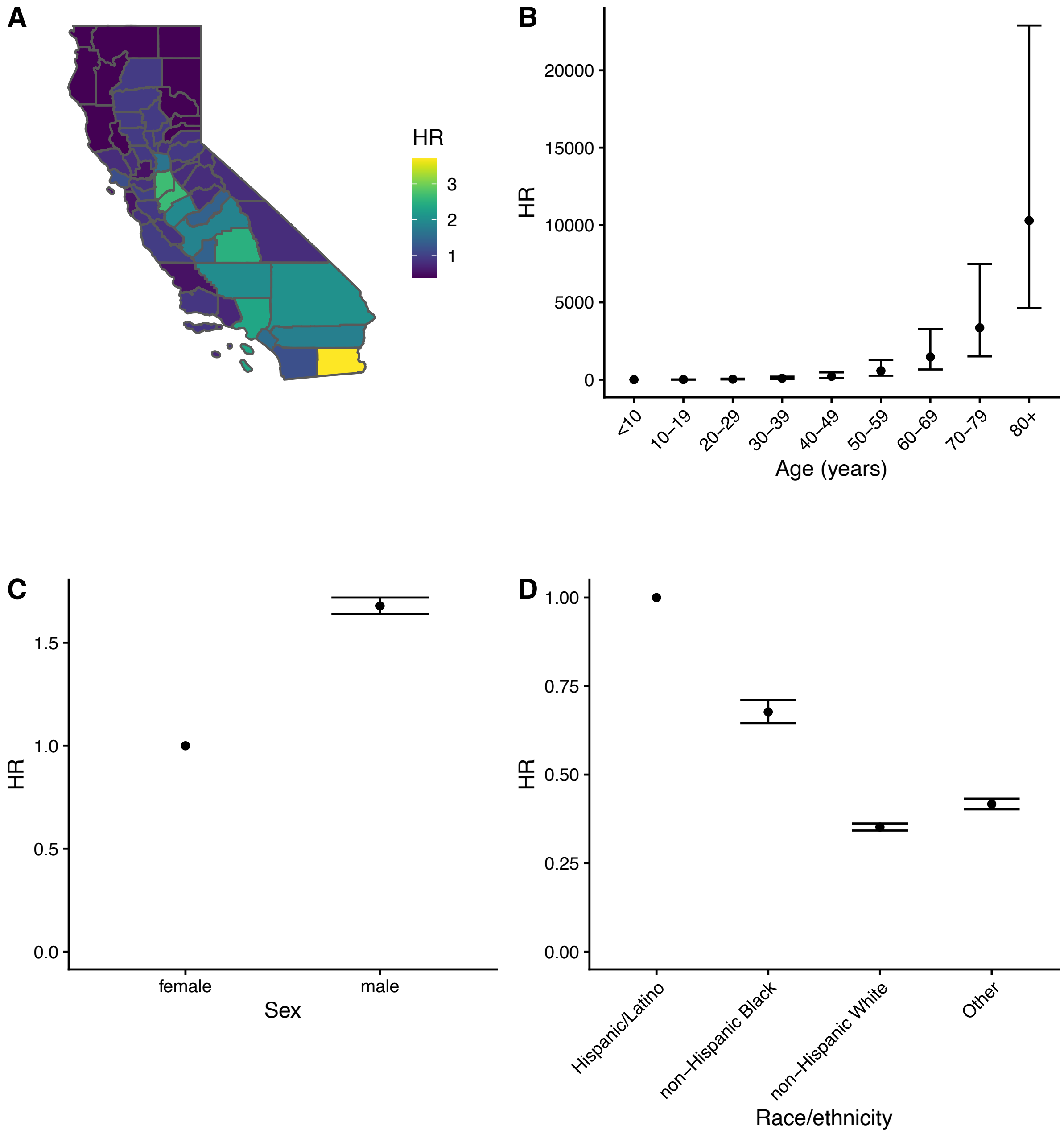


**Figure S2. Estimates of hazard ratios for risk of death from COVID-19 for demographic risk factors.** Hazard ratios (HR) for (A) county of residence, (B) age, (C) sex, and (D) race/ethnicity. Estimates from Poisson regression model fitted to data on 28,175 COVID-19 deaths in California from February 5, 2020 to December 30, 2020, from the California Department of Public Health. Counties with population <250,000 (except Imperial) were combined into a single region by their economic region, San Benito County was combined with Santa Cruz County, and Napa County was combined with Sonoma County. Plotted hazard ratios for these counties represent the hazard ratio of the combined region.

**Table S6. Sensitivity of COVID-19 death risk model parameter estimates to time period of data used.**

| Parameter | Maximum likelihood estimate (95% CI) | | |
|---|---|---|---|
| | Feb 5, 2020 - Dec 30,2020 | Jun 30, 2020 - Dec 30,2020 | Sep 30, 2020 - Dec 30, 2020 |
| Baseline COVID-19 death hazard rate (deaths/million person-days) | 0.0023 (0.001-0.0051) | 0.0042 (0.0019-0.0094) | 0.0036 (0.0012-0.011) |
| HRs for counties | | | |
| Alameda | 1 (Ref.) | 1 (Ref.) | 1 (Ref.) |
| Central Sierra | 0.8 (0.63-1) | 1 (0.82-1.3) | 0.61 (0.39-0.95) |
| Contra Costa | 0.85 (0.75-0.96) | 0.9 (0.78-1) | 0.85 (0.69-1.1) |
| Fresno | 1.9 (1.7-2.1) | 2.2 (1.9-2.4) | 2.2 (1.8-2.6) |
| Greater Sacramento | 0.93 (0.79-1.1) | 1.1 (0.89-1.2) | 1.7 (1.4-2.2) |
| Imperial | 3.7 (3.3-4.2) | 2.8 (2.4-3.3) | 2.2 (1.7-2.8) |
| Kern | 2 (1.8-2.2) | 2.2 (2-2.5) | 1.9 (1.6-2.3) |
| Los Angeles | 2.4 (2.2-2.5) | 2.1 (1.9-2.3) | 2.7 (2.3-3.1) |
| Marin | 1.2 (0.98-1.4) | 1.3 (1.1-1.6) | 0.51 (0.33-0.8) |
| Merced | 1.9 (1.7-2.3) | 2.4 (2-2.8) | 2.3 (1.8-3) |
| Monterey | 0.97 (0.82-1.1) | 1.2 (0.98-1.4) | 1.8 (1.4-2.3) |
| Napa-Sonoma | 0.79 (0.68-0.92) | 0.98 (0.84-1.2) | 0.83 (0.64-1.1) |
| Northern California | 0.38 (0.3-0.47) | 0.47 (0.38-0.6) | 0.56 (0.41-0.78) |
| Northern Sacramento Valley | 0.95 (0.81-1.1) | 1.2 (1-1.4) | 1.7 (1.3-2.1) |
| Orange | 1.6 (1.4-1.7) | 1.6 (1.4-1.7) | 1.4 (1.2-1.6) |
| Placer | 0.83 (0.69-1) | 1 (0.83-1.2) | 1.4 (1.1-1.9) |
| Riverside | 1.8 (1.6-2) | 1.7 (1.6-1.9) | 1.9 (1.7-2.3) |
| Sacramento | 1.7 (1.5-1.8) | 2 (1.8-2.2) | 2.3 (2-2.8) |
| San Bernardino | 2.1 (1.9-2.2) | 2.1 (1.9-2.4) | 1.8 (1.6-2.2) |
| San Diego | 1.2 (1.1-1.3) | 1.2 (1.1-1.3) | 1.8 (1.5-2.1) |
| San Francisco | 0.55 (0.47-0.65) | 0.54 (0.45-0.65) | 0.57 (0.44-0.75) |
| San Joaquin | 2.7 (2.4-3) | 3.2 (2.8-3.6) | 2.1 (1.7-2.6) |
| San Joaquin Valley | 1.4 (1.2-1.7) | 1.4 (1.2-1.7) | 1 (0.72-1.4) |
| San Luis Obispo | 0.53 (0.41-0.68) | 0.68 (0.52-0.88) | 1 (0.72-1.4) |
| San Mateo | 0.53 (0.45-0.63) | 0.25 (0.19-0.32) | 0.24 (0.16-0.36) |
| Santa Barbara | 0.89 (0.75-1.1) | 0.95 (0.78-1.1) | 0.77 (0.57-1.1) |
| Santa Clara | 0.93 (0.83-1) | 0.92 (0.81-1) | 1.4 (1.1-1.6) |
| SantaCruz-SanBenito | 0.84 (0.69-1) | 1 (0.84-1.3) | 2 (1.6-2.6) |
| Solano | 0.52 (0.42-0.64) | 0.51 (0.4-0.65) | 0.3 (0.19-0.49) |
| Stanislaus | 2.6 (2.4-2.9) | 3.1 (2.7-3.5) | 2.4 (2-3) |
| Tulare | 2.5 (2.2-2.8) | 2.3 (2-2.7) | 2.8 (2.3-3.5) |
| Ventura | 0.73 (0.63-0.84) | 0.77 (0.66-0.9) | 0.84 (0.66-1.1) |
| HRs for age | | | |
| <10 | 1 (Ref.) | 1 (Ref.) | 1 (Ref.) |
| 10-19 | 6.5 (2.8-15) | 5.9 (2.5-14) | 5.5 (1.6-19) |

| | | | |
|---|---|---|---|
| 20-29 | 30 (13-67) | 25 (11-57) | 17 (5.5-56) |
| 30-39 | 88 (39-200) | 65 (29-140) | 55 (17-170) |
| 40-49 | 210 (94-470) | 160 (74-370) | 140 (45-430) |
| 50-59 | 580 (260-1300) | 450 (200-1000) | 390 (120-1200) |
| 60-69 | 1500 (660-3300) | 1200 (520-2600) | 1000 (320-3100) |
| 70-79 | 3400 (1500-7500) | 2600 (1200-5700) | 2500 (790-7600) |
| 80+ | 10000 (4600-23000) | 7700 (3400-17000) | 7300 (2400-23000) |
| HR for male sex | 1.7 (1.6-1.7) | 1.7 (1.6-1.7) | 1.7 (1.6-1.8) |
| HRs for race/ethnicity | | | |
| Hispanic/Latino | 1 (Ref.) | 1 (Ref.) | 1 (Ref.) |
| non-Hispanic Black | 0.68 (0.65-0.71) | 0.61 (0.57-0.64) | 0.58 (0.53-0.63) |
| non-Hispanic White | 0.35 (0.34-0.36) | 0.34 (0.32-0.35) | 0.38 (0.36-0.4) |
| Other | 0.42 (0.4-0.43) | 0.4 (0.39-0.42) | 0.47 (0.45-0.5) |

Parameter estimates for COVID-19 death risk regression model for different past periods of data on deaths (~11 months from date of first recorded COVID-19 death to end of data, 6 months, and 3 months) from the California Department of Public Health. The estimates derived from ~11 months of data were used in the simulation of the different vaccine prioritization strategies.

**Table S7. Simulated impact of different COVID-19 vaccine prioritization strategies in California on numbers of averted COVID-19 clinical cases, deaths and DALYs for 2 million individuals vaccinated.**

| Strategy | Cases averted, mean (95% UI) | Percentage of cases averted, mean (95% UI) | Deaths averted, mean (95% UI) | Percentage of deaths averted, mean (95% UI) | DALYs averted, mean (95% UI) | Percentage of DALYs averted, mean (95% UI) |
|---|---|---|---|---|---|---|
| i) Random allocation | 38000 (28000-48000) | 6 (5-8) | 3900 (3800-4100) | 28 (27-29) | 37000 (35000-38000) | 17 (17-18) |
| ii) Special population targeting | 40000 (29000-50000) | 6 (5-8) | 3900 (3800-4000) | 28 (27-28) | 37000 (35000-39000) | 18 (17-18) |
| iii) Age targeting | 37000 (28000-46000) | 6 (4-8) | 4300 (4100-4500) | 31 (30-32) | 39000 (37000-40000) | 18 (18-19) |
| iv) Essential worker targeting | 38000 (29000-48000) | 6 (5-8) | 3900 (3700-4000) | 28 (27-28) | 36000 (35000-38000) | 17 (17-18) |
| v) Comorbidity targeting | 38000 (29000-47000) | 6 (5-8) | 3900 (3800-4100) | 28 (27-29) | 37000 (35000-39000) | 18 (17-18) |
| vi) Age-and-county targeting | 38000 (29000-47000) | 6 (5-8) | 4600 (4400-4800) | 33 (32-34) | 41000 (39000-43000) | 19 (19-20) |
| vii) Age-and-special-population targeting | 37000 (28000-47000) | 6 (4-8) | 4300 (4100-4400) | 30 (29-31) | 39000 (37000-40000) | 18 (18-19) |
| viii) Optimal allocation | 43000 (34000-52000) | 7 (5-9) | 5000 (4900-5200) | 35 (35-36) | 48000 (46000-49000) | 23 (22-23) |

Vaccine impact simulated over 6 months. All strategies assumed that all healthcare workers and long-term care facility residents were vaccinated first as per the CDC recommendation (44).

**Table S8. Simulated impact of different COVID-19 vaccination prioritization strategies in California on numbers of averted COVID-19 clinical cases, deaths and DALYs for 10 million individuals vaccinated**

| Strategy | Cases averted, mean (95% UI) | Percentage of cases averted, mean (95% UI) | Deaths averted, mean (95% UI) | Percentage of deaths averted, mean (95% UI) | DALYs averted, mean (95% UI) | Percentage of DALYs averted, mean (95% UI) |
|---|---|---|---|---|---|---|
| i) Random allocation | 150000 (130000-170000) | 24 (20-27) | 6500 (6300-6800) | 46 (45-48) | 82000 (77000-85000) | 39 (37-40) |
| ii) Special population targeting | 140000 (130000-170000) | 24 (20-27) | 6400 (6100-6700) | 46 (44-47) | 80000 (77000-84000) | 38 (37-40) |
| iii) Age targeting | 130000 (120000-150000) | 22 (19-24) | 11000 (11000-11000) | 79 (79-80) | 130000 (130000-130000) | 62 (61-63) |
| iv) Essential worker targeting | 160000 (140000-180000) | 26 (22-29) | 5300 (5100-5400) | 37 (37-38) | 74000 (71000-77000) | 35 (34-36) |
| v) Comorbidity targeting | 150000 (140000-170000) | 25 (22-29) | 7600 (7400-7900) | 54 (53-56) | 96000 (93000-1e+05) | 46 (44-47) |
| vi) Age-and-county targeting | 170000 (150000-180000) | 27 (24-30) | 11000 (11000-12000) | 81 (80-82) | 140000 (140000-150000) | 68 (67-69) |
| vii) Age-and-special-population targeting | 140000 (120000-150000) | 22 (20-25) | 11000 (11000-11000) | 80 (79-80) | 130000 (130000-140000) | 63 (62-64) |
| viii) Optimal allocation | 2e+05 (190000-220000) | 33 (30-36) | 11000 (11000-12000) | 81 (81-82) | 150000 (150000-150000) | 72 (71-73) |

Vaccine impact simulated over 6 months. All strategies assumed that all healthcare workers and long-term care facility residents were vaccinated first as per the CDC recommendation (44).

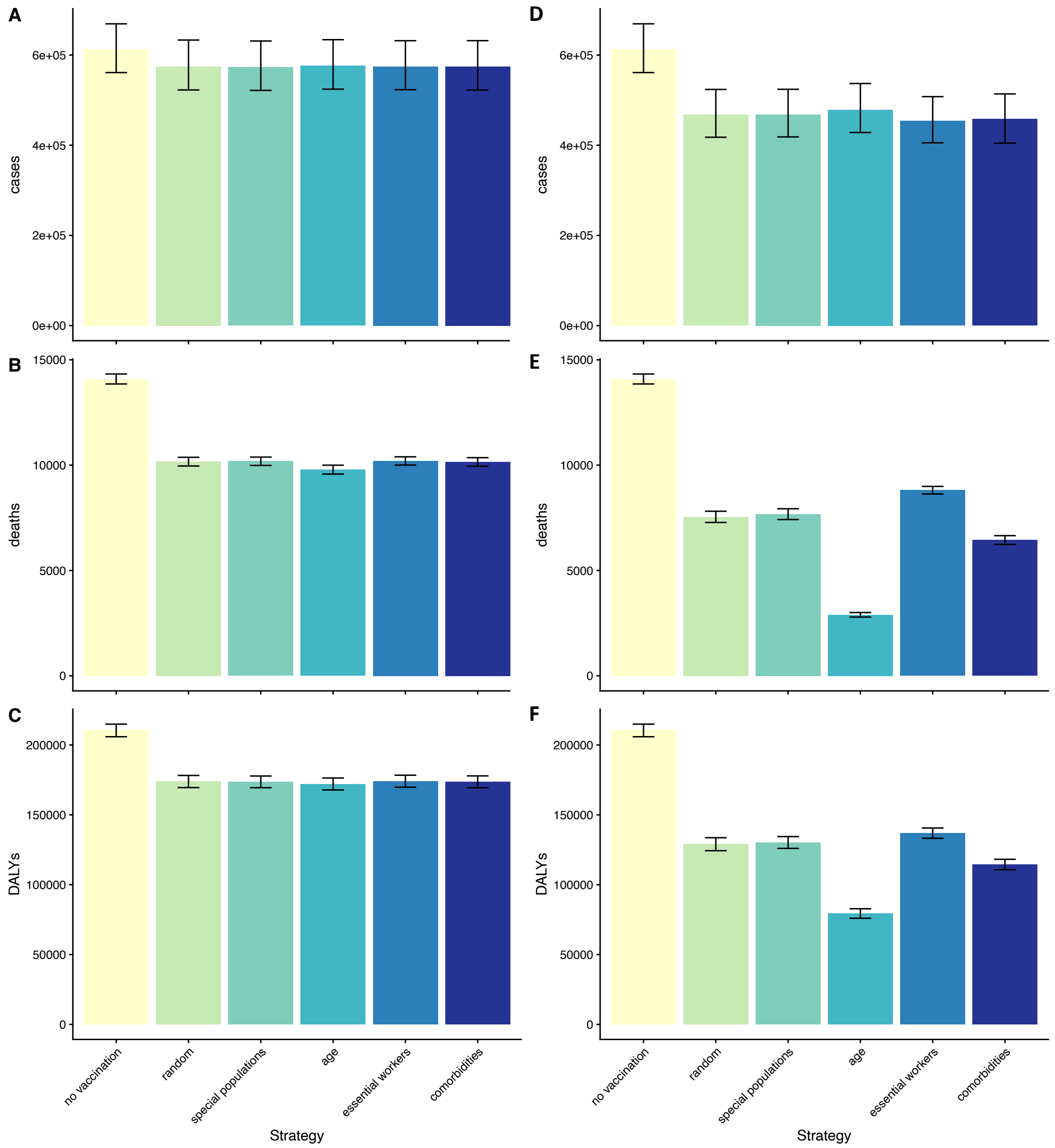


**Figure S3. Simulated impact of different COVID-19 vaccine prioritization strategies in California on numbers of COVID-19 clinical cases, deaths and DALYs for 2 million (left column) and 10 million (right column) individuals vaccinated.** Simulated (A-B) clinical cases, (C-D) deaths and (E-F) DALYs in California in the first 6 months of 2021. Vaccine efficacies against clinical disease and death were assumed to be initially 90% and 95% across all ages and to wane gradually over time (see *Simulation of COVD-19 deaths* and *Estimation of infections and clinical cases*). Error bars show 95% uncertainty intervals from stochastic uncertainty and parameter uncertainty.

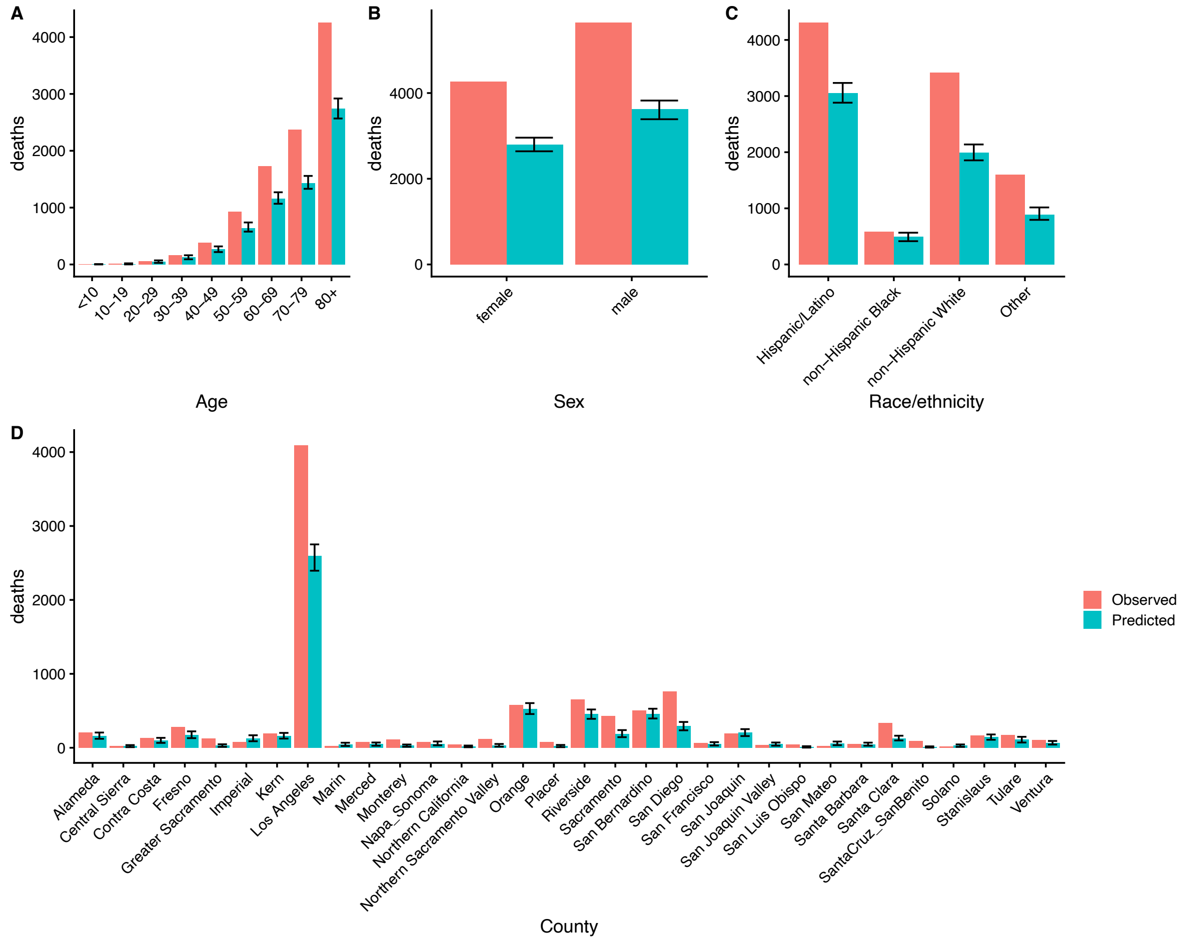


**Figure S4. Model checking.** Observed COVID-19 deaths in California Department of Public Health data and predicted COVID-19 deaths from the model for September 30 - December 30, 2020, by (A) age, (B) sex, (C) race/ethnicity and (D) county of residence. Model was fitted to data on 17,671 COVID-19 deaths from February 5 - September 30, 2020, and then simulated 1000 times to predict deaths from October 1 - December 30, 2020. Bars for predicted deaths represent mean of 1000 simulations, and error bars show 95% uncertainty intervals of the model predictions.

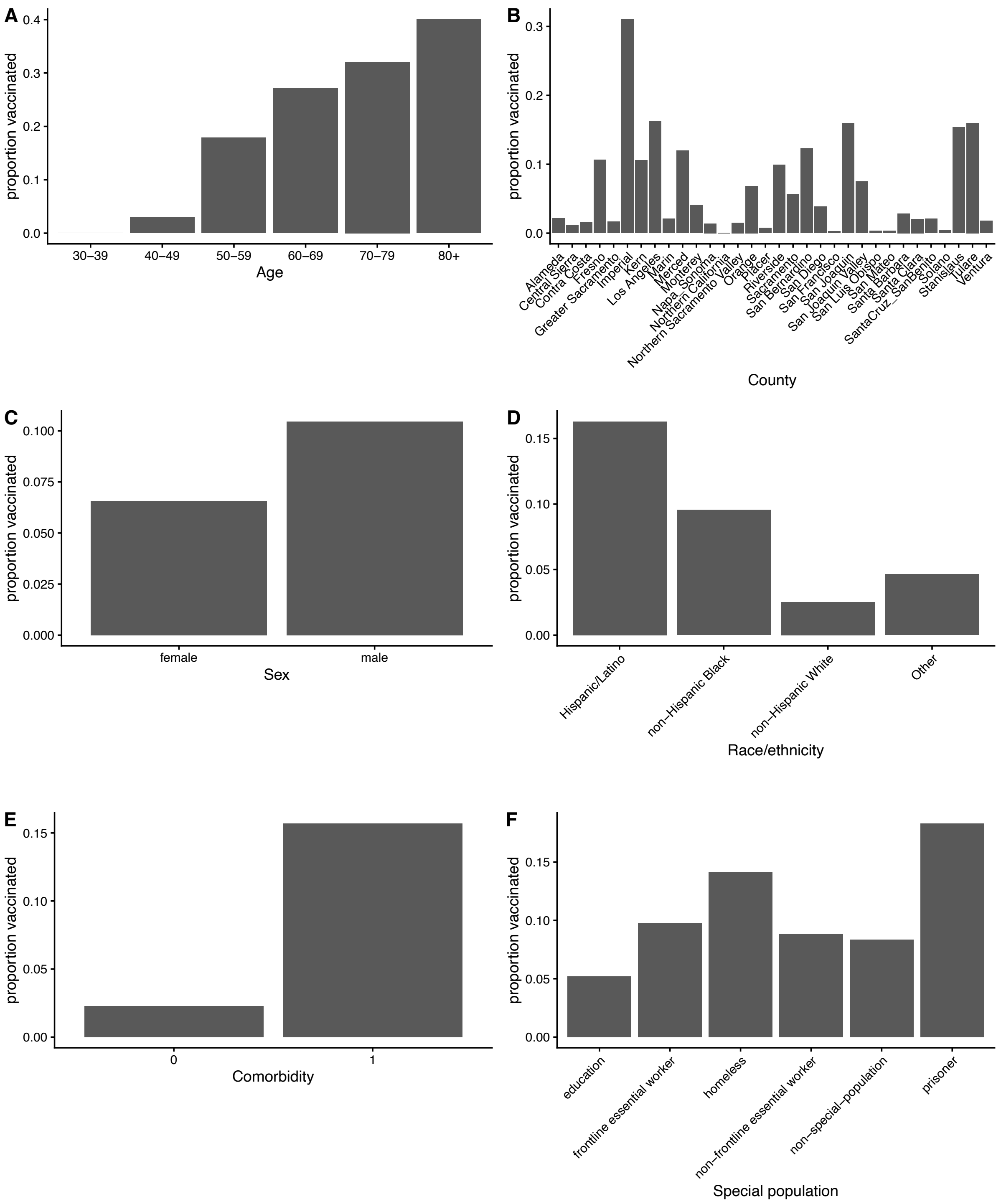


**Figure S5. Optimal vaccine allocation under simultaneous targeting by multiple risk factors.** Plots show proportion of individuals of each (A) age group, (B) county, (C) sex, (D) race/ethnicity, (E) comorbidity status, and (F) special population vaccinated under optimal distribution of the first 5 million vaccinations in California to avert DALYs. Comorbidity status treated as binary (0 = no comorbidities, 1 = any comorbidities).

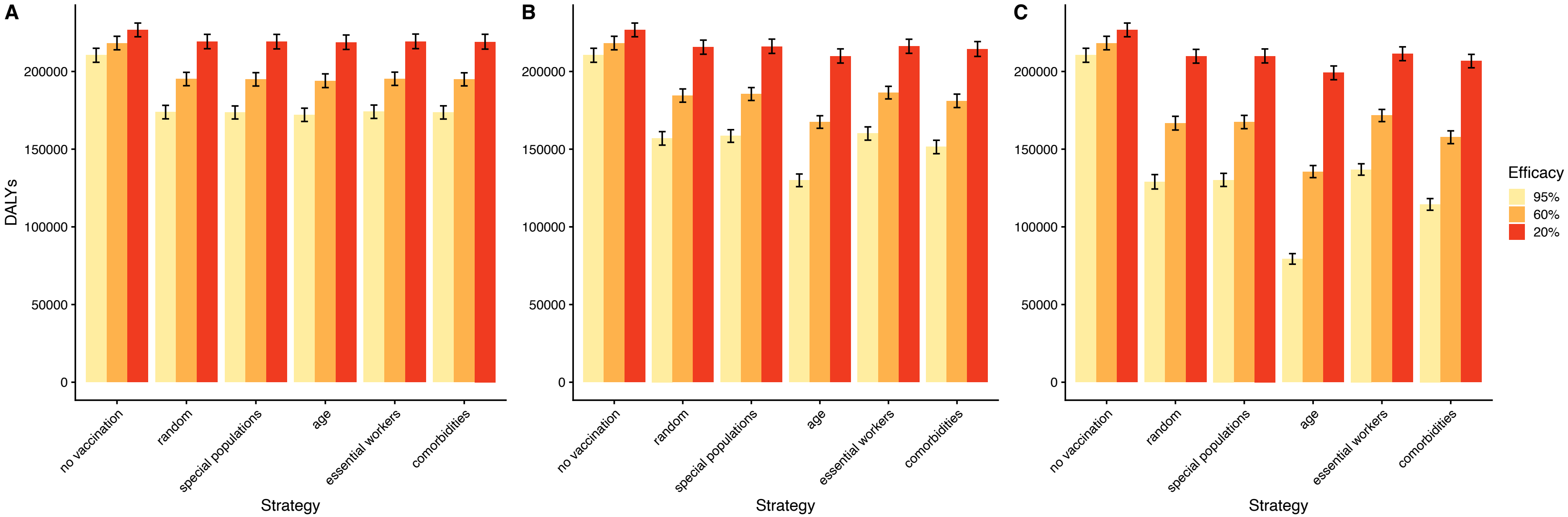


**Figure S6. Sensitivity of impact of COVID-19 vaccine prioritization strategies to vaccine efficacy.** Simulated impact of different prioritization strategies on total DALYs in California over 6 months for different vaccine efficacies (95%, 60%, and 20% across all ages) for (A) 2 million, (B) 5 million and (C) 10 million vaccinations. Error bars show 95% uncertainty intervals from stochastic uncertainty and parameter uncertainty.

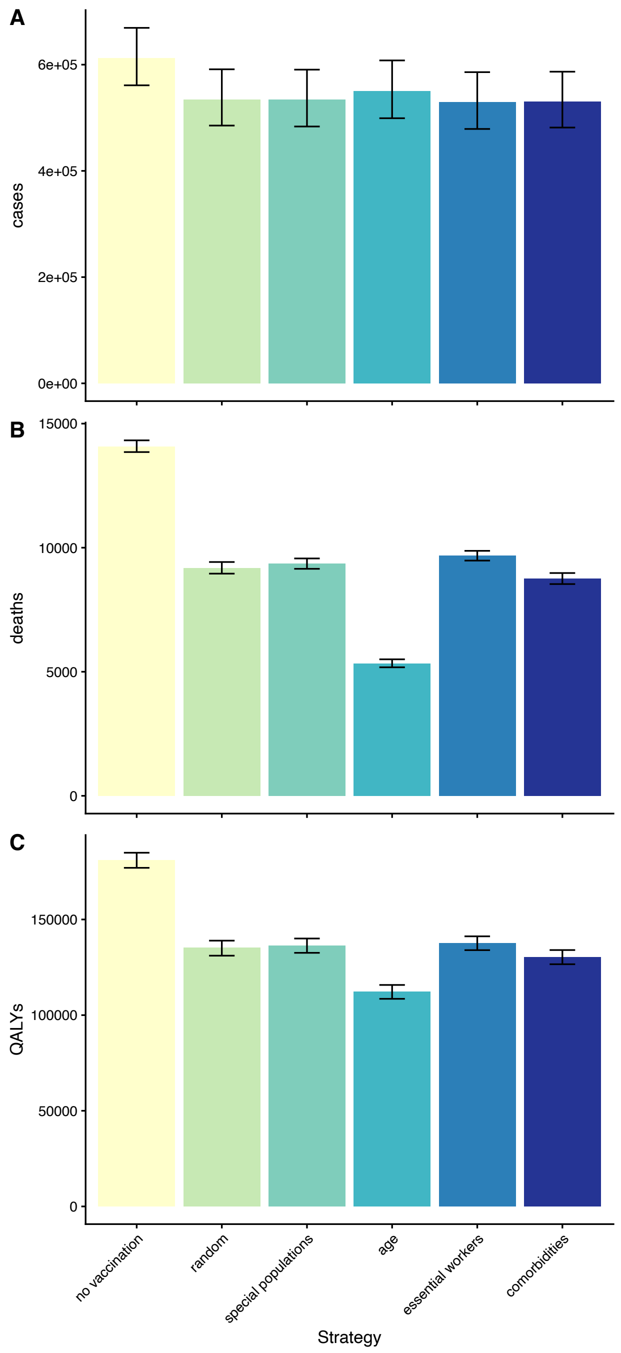


**Figure S7. Simulated impact of COVID-19 vaccine prioritization strategies in California on numbers of clinical cases, deaths and QALYs when vaccinations are allocated within each strategy according to risk of QALY loss rather than risk of DALY burden.** Simulated (A) clinical cases, (B) deaths and (C) QALYs in California in the first 6 months of 2021 with 5 million vaccinations. Vaccine efficacies against clinical disease and death as in Figure 3 in the main text. Bars show mean estimates across 1000 simulations, error bars show 95% uncertainty intervals from stochastic uncertainty and parameter uncertainty.

**Table S9. Simulated impact of different COVID-19 vaccination prioritization strategies in California on numbers of averted COVID-19 clinical cases, deaths and QALYs for 5 million individuals vaccinated when vaccinations are allocated within each strategy according to risk of QALY loss rather than risk of DALY burden**

| Strategy | Cases averted, mean (95% UI) | Percentage of cases averted, mean (95% UI) | Deaths averted, mean (95% UI) | Percentage of deaths averted, mean (95% UI) | QALYs averted, mean (95% UI) | Percentage of QALYs averted, mean (95% UI) |
|---|---|---|---|---|---|---|
| i) Random allocation | 78000 (64000-94000) | 13 (10-15) | 4900 (4700-5100) | 35 (33-36) | 46000 (44000-48000) | 25 (24-27) |
| ii) Special population targeting | 78000 (64000-93000) | 13 (10-15) | 4700 (4500-4900) | 34 (32-35) | 45000 (43000-47000) | 25 (24-26) |
| iii) Age targeting | 62000 (53000-72000) | 10 (8-12) | 8800 (8500-9000) | 62 (61-63) | 69000 (67000-71000) | 38 (37-39) |
| iv) Essential worker targeting | 84000 (68000-100000) | 14 (11-16) | 4400 (4200-4600) | 31 (30-32) | 43000 (41000-45000) | 24 (23-25) |
| v) Comorbidity targeting | 82000 (69000-96000) | 13 (11-16) | 5300 (5100-5500) | 38 (37-39) | 51000 (48000-53000) | 28 (27-29) |
| vi) Age-and-county targeting | 89000 (79000-99000) | 15 (13-17) | 9100 (8900-9300) | 64 (63-65) | 82000 (80000-84000) | 45 (44-46) |
| vii) Age-and-special-population targeting | 63000 (53000-72000) | 10 (9-12) | 8800 (8600-9000) | 62 (61-63) | 69000 (67000-71000) | 38 (37-39) |
| viii) Optimal allocation | 120000 (110000-130000) | 20 (17-22) | 9200 (9000-9400) | 65 (64-66) | 94000 (92000-97000) | 52 (51-53) |

Vaccine impact simulated over 6 months. All strategies assumed that all healthcare workers and long-term care facility residents were vaccinated first as per the CDC recommendation (44).

**Table S10. Infection fatality rate estimates from** (9) **recalibrated to California**

| | **Infection fatality rate (%)** | |
|---|---|---|
| **Age group (years)** | **Female** | **Male** |
| <10 | 0.005 | 0.005 |
| 10-19 | 0.004 | 0.005 |
| 20-29 | 0.01 | 0.03 |
| 30-39 | 0.04 | 0.12 |
| 40-49 | 0.15 | 0.36 |
| 50-59 | 0.42 | 0.98 |
| 60-69 | 1.34 | 2.70 |
| 70-79 | 4.21 | 8.67 |
| 80+ | 15.2 | 28.6 |

Median ensemble infection fatality rate estimates from (9) recalibrated for California to match cumulative number of observed cases in California Department of Public Health data (see *Calibration of infection fatality rate for California*).
